# The Burden and Impact of Intervention Strategies for Heart Failure in China (2025–2035): A Health-Augmented Macroeconomic Modeling Study

**DOI:** 10.1101/2025.04.04.25325219

**Authors:** Hua Wang, Huiyi Zhou, Ke Chai, Shaohua Yan, Yujia Liu, Michael Kuhn, Klaus Prettner, Zhong Cao, Minghui Du, Ting Wang, Ping Zeng, Jing Wu, Simiao Chen, Jiefu Yang

## Abstract

**Background:** Heart failure (HF) imposes a growing public health and macroeconomic burden in low- and middle-income countries (LMICs), yet its long-term economic implications remain unquantified. China, characterized by rapid population aging and escalating cardiovascular risks, provides a critical setting to model HF’s economic impact.

**Methods:** Using data from the Global Burden of Disease Study 2021, China Cardiovascular Association Registry, and national insurance databases, we quantified the macroeconomic burden of heart failure (2025–2035) through a health-augmented macroeconomic model. We evaluated three interventions: BNP screening (adults ≥40 years), intensive blood pressure control (hypertensive patients), and guideline-directed medical therapy (GDMT) for HF with reduced ejection fraction (HFrEF). Costs are reported in 2017 international dollars (INT$).

**Findings:** By 2035, HF cases in China are projected to reach 22·7 million (95% UI: 9·5–36·9 million), with an age-standardized prevalence of 760·65 per 100,000 (283·21–1,340·77). The cumulative economic burden (2025–2035) is estimated at INT$1,001·1 billion (733·4–1,346·6 billion), representing 0·256%(0·188%–0·344%) of annual GDP, driven by labor force attrition (72·1%; 64·4–74·8%) and direct medical costs (27·9%; 25·2–35·6%). Three interventions could reduce the total burden by 12·5%: BNP screening (25% coverage among adults≥40 years) could save INT$78·5 billion (62·8–94·1 billion; 8·10% burden reduction; cost-benefit ratio= 0·49; 0·39–0·59); intensive blood pressure control (41·4% coverage among hypertensive patients) could reduce costs by INT$27·5 billion (25·1–29·9 billion; 2·74% reduction; ratio=0·22; 0·20–0·24); and GDMT for incident HFrEF patients could yield savings of INT$17·0 billion (12·8–22·4 billion; 1·70% reduction; ratio=0·48; 0·36–0·63).

**Interpretation:** This study highlights HF’s dual clinical and macroeconomic burden in China, advocating three scalable strategies: nationwide BNP screening in primary care, subsidized hypertension management, and GDMT optimization. These interventions might offer a blueprint for LMICs to mitigate HF-related economic losses amid demographic aging.

**Funding:** This study was supported by grants from the Noncommunicable Chronic Diseases-National Science and Technology Major Project (No: 2023ZD0504600); Capital’s Funds for Health Improvement and Research (2022-1-4052); the National High-Level Hospital Clinical Research Funding (BJYY-2023-070); the National Natural Science Foundation of China (No. 82170396); and the CAMS Innovation Fund for Medical Sciences (2021-I2M-1-050).

**Research in Context:** *Evidence Before This Study:* We systematically searched MEDLINE, PubMed, and Google Scholar for studies published between 1990–2025 using terms including “heart failure,” “economic burden,” “economic cost.” Existing literature predominantly focused on cost-of-illness analyses in high-income countries, aggregating direct medical and indirect productivity costs. While the Global Burden of Disease Study provided epidemiological estimates, no prior study quantified the macroeconomic impact of heart failure (HF) in rapidly aging populations like China. Evidence on cost-effective interventions tailored to China’s healthcare systems, such as biomarker screening or workforce-targeted therapies, remained sparse.

*Added Value of This Study:* This study presents the first macroeconomic projections of HF in China, forecasting that by 2035, the number of HF cases will reach 22·7 million (uncertainty interval [UI]: 9·5 – 36·9 million), with an age-standardized prevalence rate of 760·65 (UI: 283·2–1,340·8) per 100,000 people. Age-specific analyses reveal a steep rise in HF prevalence across all age groups in China, with the most marked increases among older adults: the 60–64 years cohort is projected to rise from 0·974% (711,347 cases) in 2021 to 1·269% (1·39 million cases) by 2035 (30·3% prevalence increase; 95·5% case surge), while younger populations (35 – 39 years) show a 14·9% relative rise, underscoring the urgent need for life-course prevention strategies to address accelerating burdens in aging and younger demographics.The total economic burden from 2025 to 2035 is projected at INT$1,001·1 billion (UI: 733·4–1,345·6 billion, at constant 2017 prices), which accounts for 0·256% (UI: 0·188%–0·344%) of China’s GDP annually. This burden is driven predominantly by indirect costs (72·1%), mainly resulting from reduced labor force participation and productivity losses, with direct medical costs contributing 27·9%.Three cost-effective interventions—BNP screening (25% coverage among adults ≥40 years; 8·10% burden reduction), intensive blood pressure control (41·4% coverage in hypertensive patients; 2·74% reduction), and guideline-directed medical therapy (GDMT) for HFrEF patients (1·70% reduction)—could collectively reduce the total economic burden by 12·5%, saving up to INT$123·0 billion (UI: 100·65 – 146·39billion) over a decade, with favorable cost-benefit ratios (0·221 – 0·489) underscoring scalability in resource-limited settings.

*Implications of All the Available Evidence:* The escalating burden of HF in China reflects challenges faced by low- and middle-income countries (LMICs) navigating demographic aging and epidemiological shifts, underscoring the imperative to integrate HF prevention into national health agendas. Our findings align with China’s Healthy China 2030 initiative, the United Nations Sustainable Development Goals and Global health 2050, advocating for policies that prioritize prevention, early detection (e.g., BNP screening), and optimized chronic disease management (e.g., GDMT) to enhance workforce sustainability. Cost-effective interventions such as BNP screening and GDMT optimization are scalable across LMICs with limited resources, offering pathways to mitigate long-term societal and economic impacts. The COVID-19 pandemic has further exposed vulnerabilities in low-income and rural healthcare systems, necessitating targeted investments in digital health technologies and infrastructure to address care disparities. Policymakers must prioritize data-driven strategies combating rural-urban gaps in access, strengthening primary care capacity, and leveraging innovation to ensure equitable and sustainable HF management in aging populations globally.

## Introduction

Heart failure (HF), the end-stage manifestation of cardiovascular diseases (CVDs), threatens global health systems and economic stability, with over 64 million affected individuals worldwide–a number projected to rise sharply due to aging populations and improved survival post-acute cardiac events.^1,2^ China exemplifies this crisis, home to 245 million hypertensive adults and 140 million diabetic patients, where HF prevalence has doubled since 2000 to 12·1 million cases.^3,4^ The rapid aging of populations and the escalating risks of CVD create critical challenges that could undermine labor productivity and overall macroeconomic growth.

However, the long-term economic consequences of heart failure (HF) in low- and middle-income countries (LMICs) remain poorly quantified, despite their critical implications for sustainable development. These consequences stem from two mutually reinforcing mechanisms. First, HF erodes the workforce through premature mortality and chronic disability among working-age populations, directly reducing labor productivity and economic output. Simultaneously, the financial burden of HF — whether shouldered by households, public health systems, or insurers — diverts resources from productive investments in education, infrastructure, and private enterprise. For instance, families spending heavily on hospitalizations may sacrifice savings for children’s education, while governments allocating funds to HF treatments might delay road construction or small business subsidies. Together, these dynamics threaten to entrench cycles of poverty and undermine long-term development goals.

Existing evidence on HF’s economic burden predominantly derives from high-income countries, focusing narrowly on direct medical costs (e.g., hospitalizations, medications).^2^ Such frameworks may inadequately capture LMIC contexts. While labour-intensive economies imply that health shocks like HF could affect a larger share of the workforce, the economic impact per individual may be lower due to reduced baseline productivity. For example, in rural China, where informal employment dominates, HF-related disability might reduce income less severely than in high-productivity sectors. However, rising age-standardized CVD mortality rates in LMICs (+12% in China, 2000 – 2019) signal an urgent need to quantify these trade-offs, particularly as health burdens increasingly overlap with working-age populations.^5,6^ Despite China’s unique position as the world’s second-largest economy undergoing rapid epidemiological transition, no study has modelled HF’s macroeconomic burden or evaluated scalable interventions tailored to its fragmented primary care system.

We integrate HF epidemiology with a health-augmented macroeconomic model to quantify labor force attrition and capital depletion. Using data from the Global Burden of Disease (GBD) Study, national registries, and insurance claims, we project HF prevalence (2025 – 2035) and its macroeconomic burden. Second, we assess three interventions prioritized for China’s healthcare landscape: (1) B-type natriuretic peptide (BNP) screening for early detection; (2) intensive blood pressure (BP) control (systolic BP <120 mmHg); and (3) guideline-directed medical therapy (GDMT) for heart failure with reduced ejection fraction (HFrEF).^7^ As HF incidence rises among adults, there is growing concern over its impact on economic productivity, particularly during peak working years.^8^

The urgency of addressing HF aligns with the *Lancet* Global Health 2035 Commission’s framework, which prioritizes CVDs to halve global premature mortality by 2050.^9^ Our findings provide actionable evidence to align HF management with China’s Healthy China 2030 initiative and the United Nations Sustainable Development Goals (SDG 3: Health; SDG 8: Decent Work).

## Methods

### Projection of HF Prevalence from 2025 to 2035 in China

This study modeled the projected prevalence of HF in China from 2025 to 2035 by integrating epidemiological data from the GBD Study 2021 and demographic projections from the United Nations World Population Prospects 2024.^5,6^ Age- and sex-specific HF prevalence estimates for 2021, including 95% uncertainty interval (UI), were sourced from the GBD 2021, a standardized global dataset spanning 204 countries. A Bayesian age-period-cohort (BAPC) framework was applied using the R package *BAPC*to project future trends, accounting for multiplicative effects of age, period, and cohort on disease dynamics.^10^ In essence, the age-period-cohort model, which is a logarithmic linear Poisson model, postulates that the multiplicative impact of age, period, and cohort is reflected in a Poisson distribution, and that a link function specific to the model is utilized. The 2021 HF prevalence (both case numbers and age-standardized prevalence rates [ASPRs]) served as the baseline, with sensitivity analyses evaluating annual prevalence fluctuations (±1% as reference scenarios).

### Macroeconomic Burden Model

We adapted a health-augmented macroeconomic model, previously applied to estimate the global burden of chronic obstructive pulmonary disease, cancers, and Alzheimer’s disease, to quantify the direct and indirect economic impacts of HF in China from 2025 to 2035.^11–13^ This model integrates epidemiological data with macroeconomic variables through two primary pathways:(1) Labor force effects: HF-related mortality and morbidity reduce the working-age population and diminish labor productivity. (2) Capital accumulation effects: The costs associated with HF treatment divert financial resources from savings and investment, thereby hindering physical capital accumulation.

The economic burden attributable to HF was calculated as the cumulative difference in gross domestic product (GDP) between two scenarios:(1) Status quo scenario: Projected GDP under current HF prevalence trends.(2) Counterfactual scenario: Hypothetical GDP assuming complete elimination of HF at zero cost from 2025 onward. The key modeling assumptions are as follows: HF mortality rates fixed at 2021 levels (China Cardiovascular Association [CCA] Heart Failure Centre Registry).^14^Per capita treatment costs constant at 2017 values.^3^ Individuals with HF are excluded from the labor force until recovery or death.Retirement ages are set at 60 years for men and 55 years for women, with no adjustments for potential delayed retirement policies. Further details regarding model assumptions are provided in the Supplementary Appendix 2 A, C, D, E, G, H, J.

### Data Sources

Prevalence and mortality rates: GBD 2021 and the CCA Database–Heart Failure Centre Registry.^5,14^ Economic data: World Bank’s World Development Indicators database, International Monetary Fund’s World Economic Outlook database, and the United Nations’ World Population Prospects (2024 revision). Healthcare costs: Data from the National Urban Employee Basic Medical Insurance program across six provinces in China (2017).^3^ All economic data were converted to 2017 international dollars (INT$) to ensure consistency and comparability.

### Intervention Impact Assessment

To address the escalating burden of HF in China, we assessed three evidence-based interventions using a health-augmented macroeconomic model: BNP screening, intensive BP control, and GDMT for HFrEF. These interventions are derived from the latest guidelines of the European Society of Cardiology (ESC), the American Heart Association/American College of Cardiology/Heart Failure Society of America (AHA/ACC/HFSA), and the Chinese Guideline for the Diagnosis and Management of HF in Primary Care.^15–17^ Specific adaptations have been made for practical implementation within China’s primary care system (Table 1).

**Table 1:**
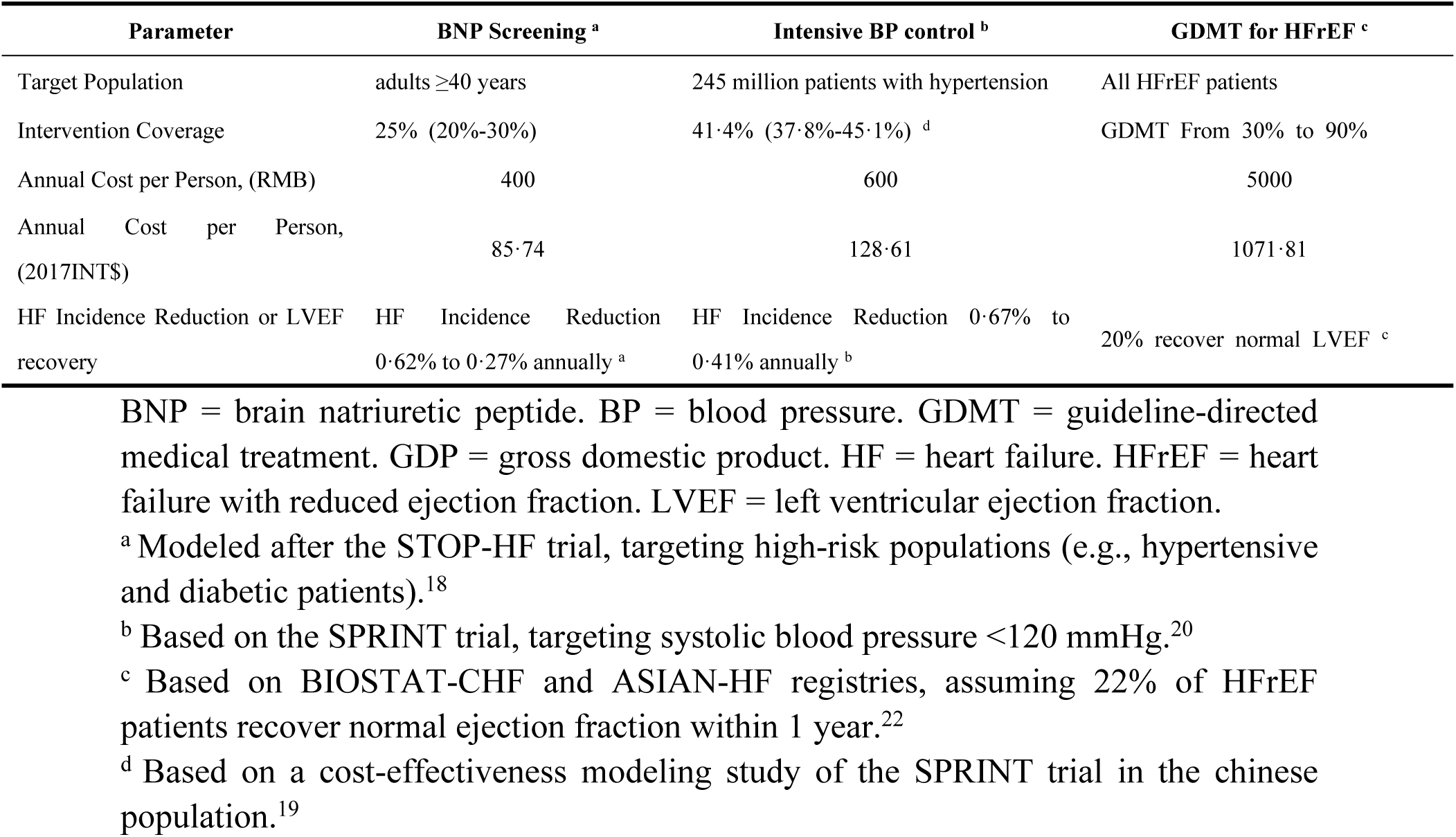
Key Input Parameters for Intervention Scenarios

### BNP Screening for Early Detection

This intervention targeted adults aged ≥40, prioritizing those with hypertension, diabetes, or a history of CVD. With an annual screening coverage of 25% (sensitivity: 20 – 30%), the cost was RMB 400 (INT$85·74) per person, covering general practitioner consultation (RMB 50), BNP testing (RMB 100), and cardiology specialist review (RMB 250). Consistent with the STOP-HF trial, BNP-based screening with collaborative care significantly reduced new-onset HF, lowering annual incidence from 0·62% to 0·27%.^18^ This effect was driven by early identification and management of at-risk individuals, preventing progression to left ventricular dysfunction.

### Intensive Blood Pressure Control

This strategy aimed to achieve a systolic BP of <120 mmHg among 245 million hypertensive adults in China, as reported in the China Hypertension Survey 2023.

Among these individuals, 41·4% (37·8 – 45·1%) met the inclusion criteria of the SPRINT trial.^19^ At an estimated annual cost of RMB 600 (INT$128·61) per patient including antihypertensive medications (RMB 350) and monitoring (RMB 250). Reduced annual HF incidence from 0·67% to 0·41%, driven by attenuated left ventricular remodeling.^20^ The China STEP trial validated this approach in older adults (target systolic BP <130 mmHg), demonstrating a 26% reduction in acute decompensated HF (HR 0·27; 95% CI 0·08–0·98).^21^

### GDMT for HFrEF

This intervention in China aimed to improve adherence to GDMT for HF patients, particularly those with HFrEF, where approximately 3 million new cases are diagnosed annually, with 35·6% being HFrEF.^3,14^ The goal was to increase adherence from 30% to 90%. The annual per-patient cost for GDMT is RMB 5,000 (2017 INT$1,071·81), which primarily covers pharmacotherapy (60 – 70%), including Sacubitril/Valsartan (RMB 1,200 – 1,800), Metoprolol (RMB 240 – 360), and Dapagliflozin (RMB 360 – 600). The remaining costs (30 – 40%) are allocated to monitoring, including BNP tests (three times per year) and echocardiographic assessments (one to two times per year). Clinically, GDMT helps 20–30% of HFrEF patients achieve left ventricular functional recovery (LVEF >40%) within 12 months, improving their physical capacity.^22^

Using the previously described health-augmented macroeconomic model, we calculated the economic burden reduction associated with each intervention. We compared annual aggregate output (GDP) across two scenarios from 2025 to 2035, the key modeling assumptions are as follows: In the status quo scenario, GDP is projected based on current estimates and disease prevalence projections. In the counterfactual scenario, HF prevalence decreases due to the intervention, while additional costs are incurred. The average duration of HF is assumed to be 4 years, annual HF incidence of 3 million cases.^3,14^ For each intervention, we estimated: Reduction in HF prevalence, Savings in healthcare expenditures, Productivity gains from reduced disability and premature mortality, Cumulative economic benefits over a 10-year horizon (2025 – 2035). Further details regarding model assumptions are provided in the Supplementary appendix 2 B.

### Sensitivity Analysis

We conducted sensitivity analyses to assess the robustness of our estimates. Baseline estimates were based on mean prevalence rates, UI were calculated in the sensitivity analysis based on the lower and upper bounds of 95% UI for GBD data.We applied discount rates of 0%, 2%, 3%, 4%, and 5%, with 3% used as the baseline in the main analysis(Supplementary appendix 2 K). Incorporated regional cost heterogeneity by calculating province-, sex-, and age-stratified treatment costs from six economically diverse provinces, using minimum and maximum values across provinces for each subgroup to evaluate cost-driven variability (Supplementary appendix 2 L). The model’s predictive accuracy was validated against historical HF epidemiology and economic burden data.

## Results

Based on data from the GBD Study 2021, the prevalence of HF in China is projected to rise substantially between 2021 and 2035, driven by rapid population aging and escalating cardiovascular risk factors (Figure 1, appendix 2 F) By 2035, the number of individuals living with HF is estimated to reach 22·7 million (95% UI: 9·496 – 36·936 million), representing a 73% increase from 2021 (13·1 million) (95% UI: 12·847 to 13·353). The age-standardized prevalence rate is expected to rise from 693·47 (95% UI: 675·18–711·75) per 100,000 in 2021 to 760·65 (95% UI: 283·21–1340·77) per 100,000 by 2035.

**Figure 1.**
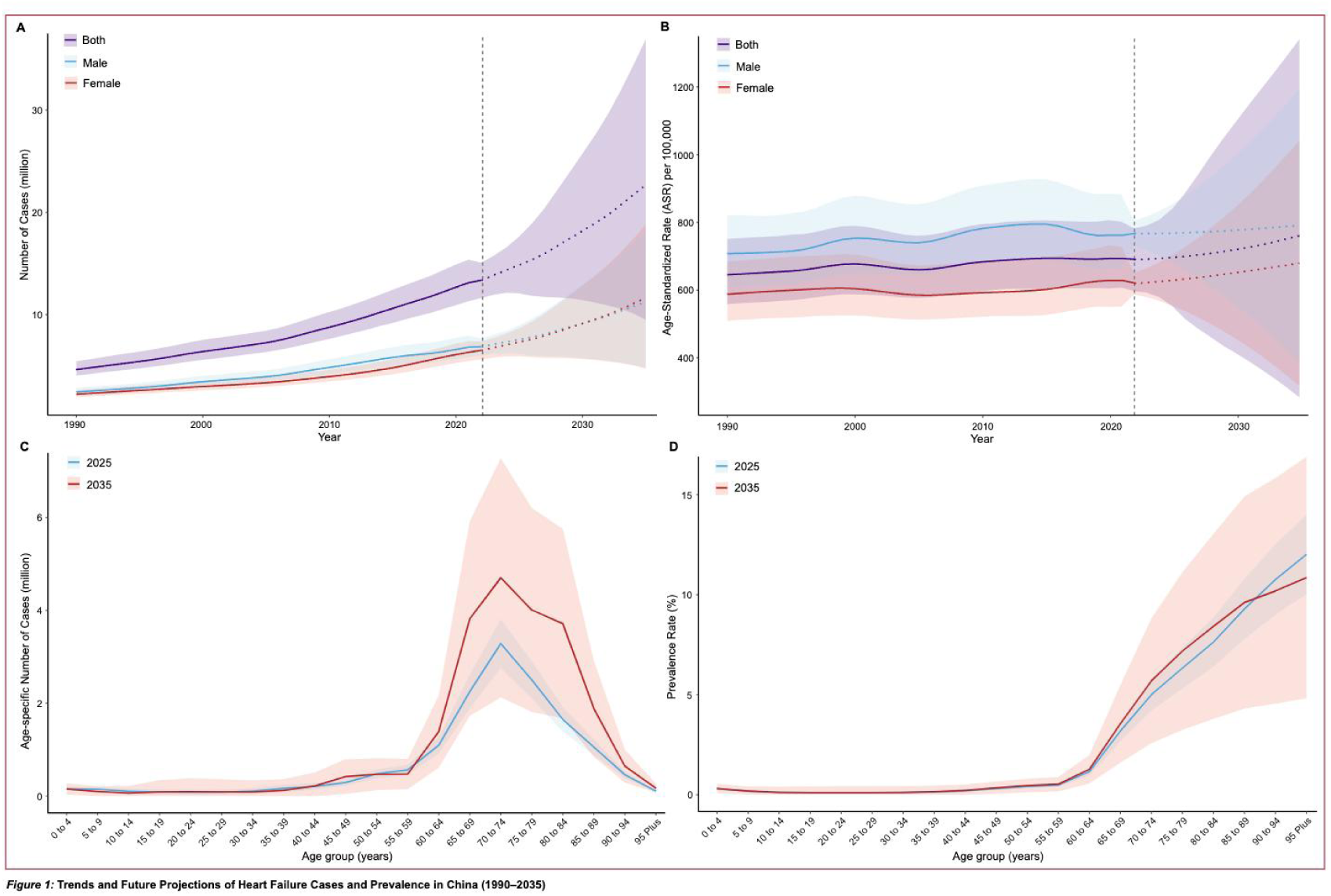
Trends and Future Projections of Heart Failure Cases and Prevalence in China (1990–2035)

Age-specific trends in HF prevalence reveal a consistent upward trajectory across all age groups in China, with the most pronounced increases observed among older populations (Figure 1, appendix 2 I). In younger adults aged 15 – 49 years, HF prevalence remains relatively low but demonstrates a gradual rise over time. For instance, the prevalence in the 35–39 years age group is projected to increase from 0·127% in 2021 to 0·146% by 2035, reflecting a 14·9% relative increase. Middle-aged populations (50–64 years) face a more substantial escalation, with the 60 – 64 years cohort anticipated to rise from 0·974% to 1·269% (a 30·3% relative increase). Cases in this group are expected to surge from 711,347 to 1·39 million (a 95·5% relative increase). The most significant burden is concentrated among older adults. Notably, the prevalence in the 65–69 years age group is projected to rise from 2·728% to 3·549% (a 30·1% relative increase), with cases surging from 2·09 million in 2021 to 3·82 million by 2035 (an 82·7% relative increase). Meanwhile, the 70–74 years cohort is expected to increase from 4·679% to 5·692% (a 21·6% relative increase), with cases rising from 2·49 million to 4·70 million (an 88·8% relative increase). Prevalence rates for males and females across various age groups can be found in Appendix 2 G, H, I.

Our macroeconomic model forecasts a significant increase in the economic burden of HF in China from 2025 to 2035 (Table 2). The burden is expected to rise from INT$46·552 billion (UI:39·563–54·418 billion) in 2025 to INT$183·512 billion (UI: 118·975 – 273·759 billion) in 2035. Concurrently, HF-related economic losses as a percentage of GDP are projected to grow from 0·137% (UI: 0.116% – 0.160%) in 2025 to 0·365% (UI: 0·2367%–0·544%) by 2035. Cumulatively, the discounted total burden over the 2025–2035 period is estimated to reach INT$1,001·141 billion (UI: 733·444 – 1,365·639 billion), representing an average annual loss of 0·256% (UI: 0.188 – 0.344%) of China’s GDP. The economic burden of HF is driven by direct medical costs (27·9%; UI: 25·2%–35·6%) and indirect costs (72·1%; UI: 64·4%–74·8%). Direct costs include hospitalizations, medications, and outpatient care, while indirect costs result from reduced labor force participation and productivity losses. Table 3 presents the impact of three interventions on reducing the economic burden of HF over a 10-year period. Among them, BNP-based screening demonstrated the highest macroeconomic efficiency, with a cumulative 10-year cost of INT$160·47 billion (UI: 128·38 –192·56 billion), leading to a reduction in economic burden by INT$78·49 billion (UI: 62·79 – 94·09 billion), equivalent to 8·10% (UI: 6·27 – 9·74%) of the total HF burden. While its impact on GDP remains marginal at 0·020% (UI: 0·016–0·024%), the intervention’s cost-benefit ratio of 0·489 (95% UI: 0·391–0·586) highlights significant returns, primarily driven by labor force retention, which accounts for 96·9% of the total benefits through early detection of at-risk populations. Intensive blood pressure control, targeting 245 million hypertensive adults, required a total investment of INT$124·33 billion (UI: 113·52 – 135·44 billion) but yielded a reduction in the HF burden by INT$27·48 billion (UI: 25·09 – 29·94 billion), contributing 2·74% (UI: 2·51 – 2·99%) of the total burden reduction with a cost-benefit ratio of 0·221 (UI: 0·202 – 0·241). While this strategy offers broader cardiovascular benefits beyond HF prevention, its scalability remains challenged by substantial upfront financial requirements, limiting its feasibility in resource-constrained settings.

**Table 2.** Projected Macroeconomic Burden of Heart Failure in China (2025–2035)

| Year | Economic Loss, Billions of 2017 INT\$ | Proportion of GDP, % | Proportion Due to Physical Capital Decline, % | Proportion Due to Labor Force Decline, % |
| --- | --- | --- | --- | --- |
| 2025 | 46·552 (39·534-54·418) | 0·137 (0·116-0·160) | 0·0 (0·0-0·0) | 100·0 (100·0-100·0) |
| 2026 | 57·595 (47·911-68·601) | 0·162 (0·135-0·193) | 9·7 (8·5-13·5) | 90·3 (86·5-91·6) |
| 2027 | 68·780 (55·959-83·590) | 0·187 (0·152-0·227) | 16·4 (14·5-22·2) | 83·6 (77·8-85·5) |
| 2028 | 80·092 (63·639-99·445) | 0·211 (0·168-0·262) | 21·4 (19·1-28·2) | 78·6 (71·8-80·9) |
| 2029 | 91·643 (71·017-116·392) | 0·234 (0·181-0·297) | 25·3 (22·7-32·7) | 74·7 (67·3-77·3) |
| 2030 | 104·662 (78·992-136·140) | 0·256 (0·193-0·333) | 28·3 (25·5-36·2) | 71·7 (63·8-74·5) |
| 2031 | 118·542 (87·022-158·109) | 0·278 (0·204-0·371) | 30·7 (27·8-38·8) | 69·3 (61·2-72·2) |
| 2032 | 133·329 (95·073-182·565) | 0·300 (0·214-0·411) | 32·7 (29·7-40·9) | 67·3 (59·1-70·3) |
| 2033 | 149·021 (103·083-209·724) | 0·322 (0·222-0·453) | 34·3 (31·3-42·6) | 65·7 (57·4-68·8) |
| 2034 | 165·660 (111·019-239·889) | 0·343 (0·230-0·497) | 35·6 (32·6-44·0) | 64·4 (56·0-67·4) |
| 2035 | 183·512 (118·975-273·759) | 0·365 (0·236-0·544) | 36·8 (33·7-45·1) | 63·2 (54·9-66·3) |
| Total (Discounted) | 1001·140 (733·444-1345·639) | 0·256 (0·188-0·344) | 27·9 (25·2-35·6) | 72·1 (64·4-74·8) |

GDP = gross domestic product.
Economic loss is expressed in billions of 2017 international dollars (2017INT\$).
The total economic burden includes direct healthcare costs (e.g., hospitalizations, medications) and indirect costs (e.g., lost productivity, disability).
Uncertainty intervals in parentheses are calculated based on the lower and upper bounds of 95% uncertainty intervals for Projected Heart Failure Prevalence by Age Group derived from the GBD Study 2021, adjusted for China-specific demographic and risk factor trends.
The uncertainty intervals for the "Proportion Due to Physical Capital Decline, %" and "Proportion Due to Labor Force Decline, %" are derived from sensitivity analyses conducted on per capita treatment costs, which were calculated by gender and age group at the provincial level using National Urban Employee Basic Medical Insurance data.
The cumulative economic burden is adjusted for discounting at a rate of 3% per year.

**Table 3.** Cost-Benefit of Heart Failure Interventions (2025–2035)

| Parameter | BNP Screening | Intensive BP control | GDMT for HFrEF |
| --- | --- | --- | --- |
| Total Cost (10-Year Cumulative), Billions of 2017 INT\$ | 160·469 (128·375-192·563) | 124·328 (113·517-135·439) | 35·327 (26·047-47·101) |
| Reduction in Patients (10-Year Cumulative) | 7,589,641 (6,071,715-9,107,573) | 2,900,898 (2,648,646-3,160,157) | 1,851,445 (1,328,668-2,526,094) |
| Economic Burden Reduction (10-Year Cumulative)<sup>a</sup>, Billions of 2017 INT\$ | 78·49 (62·79-94·09) | 27·48 (25·09-29·94) | 17·03 (12·77-22·36) |
| Relative Economic Burden Reduction Rate, % | 8·10 (6·27-9·74) | 2·74 (2·51-2·99) | 1·70 (1·66-1·74) |
| GDP Reduction, % | 0·020 (0·016-0·024) | 0·007(0·006-0·008) | 0·004 (0·003-0·006) |
| Contribution to Burden Reduction |  |  |  |
| Physical Capital Improvement<sup>b</sup>, % | 3·1 | -35·0 | 3·5 (3·2-3·9) |
| Labor Force Improvement<sup>c</sup>, % | 96·9 | 135·0<sup>e</sup> | 96·5 (96·1-96·8) |
| Cost-benefit Ratio<sup>d</sup> (Burden Reduction per Unit Cost) | 0·489 (0·391-0·586) | 0·221 (0·202-0·241) | 0·482 (0·361-0·633) |

BNP = brain natriuretic peptide. BP = blood pressure. GDMT = guideline-directed medical treatment.
GDP = gross domestic product. HF = heart failure. HFrEF = heart failure with reduced ejection fraction.
LVEF = left ventricular ejection fraction.
<sup>a</sup> Economic burden reductions are adjusted for discounting at a rate of 3% per year.
<sup>b</sup> Physical capital improvement captures savings from reduced healthcare expenditures redirected to investments; negative values indicate net capital depletion due to intervention costs.
<sup>c</sup> Labor force improvement exceeding 100% reflects indirect productivity gains from reduced caregiver burden and avoided premature mortality, which amplify direct workforce retention effects.
<sup>d</sup> Cost-benefit ratio = Economic Burden Reduction / Total Cost. Values >0.5 indicate high efficiency.

The GDMT for HFrEF has emerged as a cost-effective and balanced strategy, requiring the lowest investment (INT$35·33 billion; UI: 26·05–47·10 billion) while reducing the economic burden by INT$17·03 billion (UI: 12·77 – 22·36 billion), equivalent to 1·70% ( UI: 1·66–1·74%) of the total burden, with a cost-benefit ratio of 0·482 (UI: 0·361 – 0·633). Notably, over 96·5% (UI: 96·1 – 96·8%) of the economic benefits were driven by productivity restoration in younger patients through improved cardiac function, with an additional 3·5% (UI: 3·2–3·9%) stemming from reduced long-term care costs. Given its efficiency and targeted impact on the working-age population, GDMT represents a sustainable and scalable intervention, particularly for resource-limited settings.

Figure 2 illustrates the comparative impact of the three interventions by presenting the annual proportion of economic burden reduction attributed to each from 2025 to 2035. The interventions exhibit distinct patterns of economic burden reduction over time. BNP-based screening demonstrates the most sustained and pronounced decline in economic burden, underscoring its long-term effectiveness with a 8·10% reduction (UI: 6·27–9·74%) of the total burden. In contrast, GDMT for HFrEF, while effective, contributes a smaller proportion (1·70%; UI: 1·66–1·74%) due to its narrower target population and higher per-person costs. The three interventions are projected to collectively reduce the economic burden by 12·54% ( UI: 10·44–14·47%) from 2025 to 2035, representing savings of INT$123·00 billion (UI: 100·65–146·39 billion).

**Figure 2.**
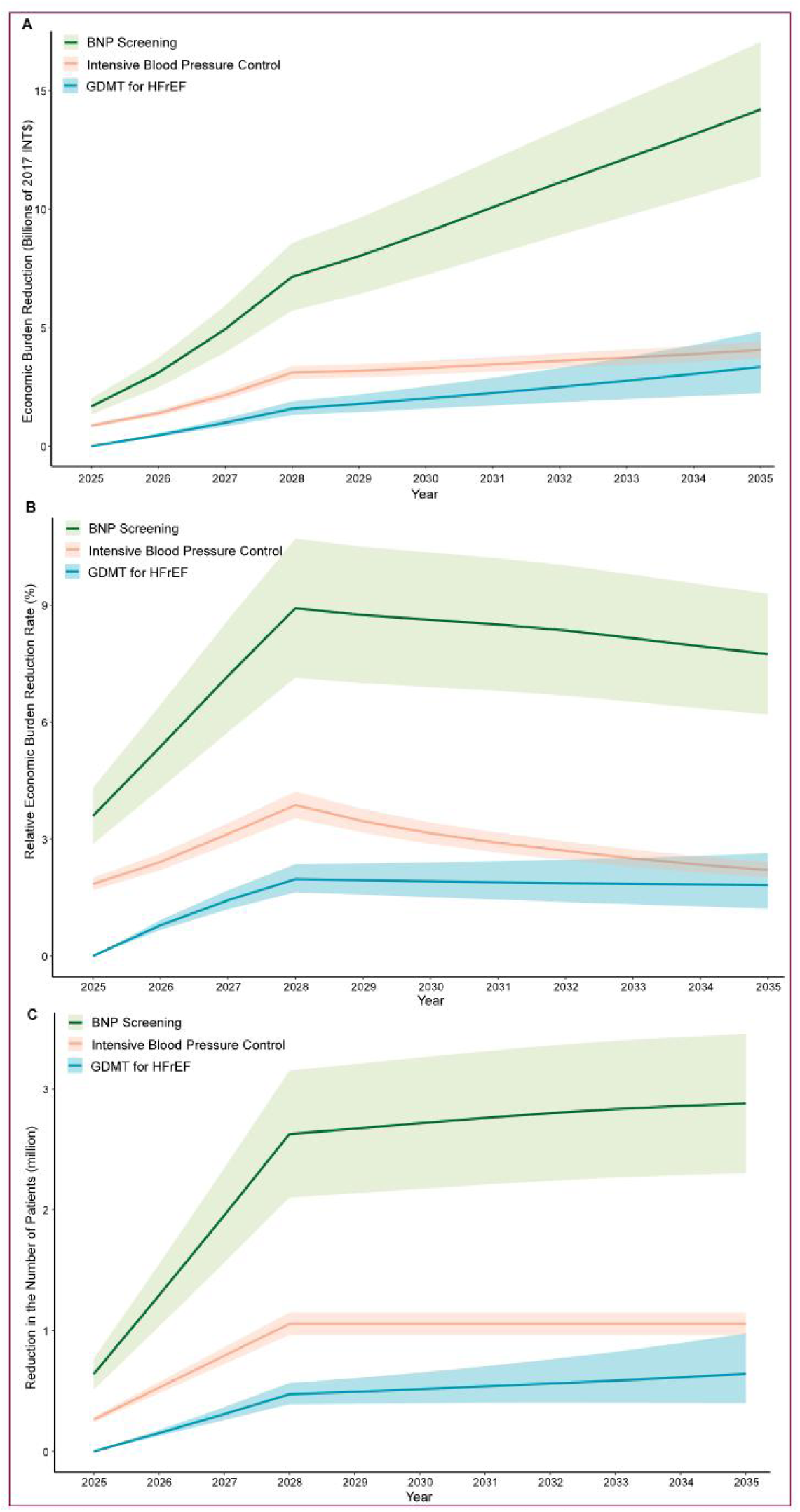
Impact of BNP Screening, Intensive Blood Pressure Control, and GDMT for HFrEF on Economic Burden and Patient Reduction (2025-2035) BNP=brain natriuretic peptide. GDMT=guideline-directed medical treatment. HF=heart failure. HFrEF=heart failure with reduced ejection fraction.

## Discussion

This study provides the first macroeconomic assessment of HF in China, projecting a cumulative economic burden of INT$1,001·1 billion (0·256% of GDP) from 2025 to 2035, driven predominantly by indirect costs from labor force attrition (72·1%). Our findings highlight three critical insights: (1) HF’s dual burden among aging and younger populations signals a looming crisis for China’s labor-driven economy; (2) scalable interventions—BNP screening, intensive BP control, and GDMT—could mitigate 12·54% of this burden (INT$123·0 billion saved); and (3) indirect costs dominate HF’s economic impact in China, contrasting sharply with high-income countries where direct medical costs prevail. These results underscore the urgency of integrating HF management into national health agendas, aligning with the Healthy China 2030 initiative and the United Nations Sustainable Development Goals (SDGs).

Globally, HF imposes escalating costs in aging populations, yet China’s economic profile amplifies its vulnerability. In high-income countries, direct medical costs dominate HF-related expenditures (65 – 80% of total burden).^2,23,24^ By contrast, China’s indirect costs (72·1%) highlight its reliance on workforce productivity. This pattern aligns with studies of chronic diseases in LMICs, such as COPD and cancer, where indirect costs similarly prevail.^12,13^ However, HF’s projected GDP loss (0·365% by 2035) surpasses that of many chronic conditions (e.g., global COPD: 0·111% GDP/year), positioning HF as a macroeconomic priority requiring systemic interventions beyond clinical management.^12^

While global age-standardized CVD prevalence remains stable, HF’s growing burden in China signals an urgent need for life-course prevention.^25^ By 2050, ischaemic heart disease is projected to cause 20 million cardiovascular deaths globally, with hypertension as the leading modifiable risk factor (18·9 million attributable deaths).^25^ Despite lower absolute HF risk in younger populations, the stronger association of modifiable risk factors (e.g., hypertension, diabetes) in adults aged 35 – 55 years demands targeted prevention strategies to mitigate productivity losses during peak working years.^26,27^

BNP-based screening emerged as the most cost-effective intervention, reducing 8·10% (INT$78·49 billion) of the total HF burden with a cost-benefit ratio of 0·49. This aligns with evidence from the STOP-HF trial, where BNP-guided care reduced left ventricular dysfunction incidence by 41% (OR 0·55; 95% CI 0·37 – 0·82).^18^Cost-effectiveness analyses support its scalability.^28^ However, successful implementation depends on integration into primary care, especially in rural China, where diagnostic tools and specialist care are limited.^29^ Scaling this intervention requires integrating BNP screening into China’s Essential Public Health Services, prioritizing adults ≥40 years in high-risk regions. Portable BNP devices paired with telemedicine platforms could bridge diagnostic gaps, while training primary care providers in risk stratification would enhance early detection.

Intensive systolic BP control could reduce HF-related economic burdens by 2·74% (INT$27·5 billion), consistent with the SPRINT trial’s 25% HF risk reduction (HR: 0·75; 95% CI: 0·64–0·89).^20^ The China STEP trial further validated this approach in older adults (target <130 mmHg), significantly lowering cardiovascular events (HR: 0·74; 95% CI: 0·60–0·92), showing a 73% decline in acute HF episodes (HR: 0·27; 95% CI: 0·08 – 0·98).^21^ To offset its 35·0% physical capital depletion (driven by medication costs), subsidizing antihypertensive therapies under national insurance is critical. Systemic barriers—including fragmented medication access and limited BP monitoring infrastructure—constrain scalability. Scalable solutions—such as village doctor-led rural programs, workplace-based BP monitoring, and digital adherence tools—aligned with China’s Essential Public Health Services framework, could bridge implementation barriers, translating clinical success into equitable health and economic outcomes.^30,31^

Optimizing GDMT for HFrEF demonstrated balanced efficiency. Despite robust evidence for ACE inhibitors, beta-blockers, and SGLT2 inhibitors, adherence remains suboptimal, with only 6·2% of patients achieving optimal GDMT use within 12 months of diagnosis.^32–35^ Significant sex disparities exist, as women receive GDMT less frequently than men.^34^ Post-discharge mortality in low-income regions correlates with inadequate GDMT prescription, reflecting systemic inequities.^36^ Scaling China’s Heart Failure Center model—which achieved 80% GDMT adherence through standardized protocols and multidisciplinary care—demonstrates actionable success.^14^ A randomized trial confirmed that digital consults improve GDMT optimization, offering scalable solutions to address fragmented access.^37^ Prioritizing equity-focused policies, multidisciplinary collaboration, AI-driven decision support tools and telehealth consultations is critical to bridging implementation gaps and translating evidence into practice.

HF poses significant clinical and macroeconomic challenges in China. Integrated policies should focus on prevention, workforce protection, and strengthening healthcare systems. Scalable interventions align with the Healthy China 2030 initiative and the Lancet Global Health 2035 Commission’s “50 by 50” goal. Despite progress, universal health coverage lags in preventive care.^29^ Investments should strengthen primary healthcare infrastructure to address the dual burden of non-communicable diseases and aging.^38^ Prioritize BNP screening and GDMT for labor force protection, as they are highly cost-effective. Implement hypertension management subsidies to mitigate short-term risks, and leverage AI and telemedicine to standardize treatment and reduce rural-urban disparities. Combining BNP screening and hypertension management could amplify benefits, reducing healthcare expenditures and improving labor force productivity. Joint interventions may lower the total burden compared to BNP screening alone. A national scale-up of BNP screening, especially in rural regions, is essential for maximum impact.

While our study advances the understanding of HF as a macroeconomic challenge, several limitations necessitate cautious interpretation of the findings. First, reliance on GBD data may obscure regional disparities in HF epidemiology and healthcare access. For example, rural-urban gaps in diagnostic capacity — such as limited echocardiography availability in remote areas — were not fully captured, potentially biasing national prevalence projections. Second, the observed wide credible intervals likely reflect compounded uncertainties from sparse subpopulation data (e.g., ethnic minorities, older adults), inherent identifiability constraints in age-period-cohort models, and insufficient smoothing priors amplifying noise in long-term forecasts. Third, fixed treatment costs exclude savings from medical innovations (e.g., SGLT2 inhibitors reducing hospitalizations by 30% in recent trials) and inflationary effects, which may underestimate long-term economic benefits of interventions.^38^ Fourth, while our analysis focused on HF-specific burdens, cross-disease cost savings from interventions— such as hypertension management reducing stroke incidence — were omitted, narrowing the estimated societal value. Fifth, uniform intervention adoption rates oversimplify implementation realities; for instance, GDMT adherence remains lower in rural versus urban China, yet our model assumed homogeneous scaling. Finally, broader socioeconomic determinants—such as air pollution and healthcare inequities—were omitted, despite evidence linking these factors to HF progression and economic vulnerability.

## Conclusion

HF in China represents not only a critical public health challenge but also imposes a substantial long-term macroeconomic burden. By prioritizing BNP screening, intensive BP control, and optimized GDMT adherence, policymakers can mitigate 12·5% of the projected burden (INT$123·0 billion saved), safeguarding GDP growth and aligning with global sustainability agendas. For China and LMICs, prioritizing HF management is an investment in both health and economic resilience.

## Contributors

HW, HZ, MK, KP, JW, SC, and JY conceptualised and designed the study. HW, HZ, SY, MD, SC, and JY acquired the data and information for this study. HW, HZ, KC, SY, YL, TW and PZ conducted the analyses, visualised and interpreted the data, and reviewed the literature. JY, JW, SC, TW, and PZ contributed to the literature review and the interpretation of the data. HW, HZ, KC, MK, KP, CZ, and SY wrote the article. JY, MK, KP, CZ, JW, SC, and HW critically revised the article. HW, HZ, JW, SC, and JY accessed and verified all the data, and they had final responsibility for the decision to submit the manuscript for publication. All authors had full access to all data used in the study and approved the final version.

## Ethics Statement

This study utilized de-identified data from three sources: the Global Burden of Disease Study 2021 (publicly available aggregate data), the China Cardiovascular Association Registry, and national insurance databases. Ethical approval for the use of the China Cardiovascular Association Registry data was obtained from the Central Ethics Committee of Beijing Hospital (Approval No.: 2022BJYYEC-346-01), as detailed in our prior publication.^14^ Access to the national insurance databases was approved by the Ethical Review Committee of Beijing Hospital (IRB No.: 2019BJYYEC-219-01), with waived informed consent due to the retrospective, anonymized nature of the data.^3^ The Global Burden of Disease Study 2021 adheres to ethical standards for global health data aggregation, ensuring no individual-level identifiers. All analyses complied with institutional and national guidelines for secondary data use, and no additional ethics review was required for this modeling study.

## Data sharing

No individual-level data were used in this modelling study. Data from this modelling study are available with publication. The data are available to anyone who requests them for any non-commercial purposes. The data can be accessed by contacting HW, who will provide guidance on how to use and interpret the data.

## Declaration of generative AI and AI-assisted technologies in the writing process

During the preparation of this work, the authors used ChatGPT and deepseek in order to improve language clarity and correct grammatical errors. After using this tool, the authors reviewed and edited the content as needed and take full responsibility for the content of the publication.

## Data Availability

All data produced in the present study are available upon reasonable request to Hua Wang

## Acknowledgements

This study was supported by grants from the Noncommunicable Chronic Diseases-National Science and Technology Major Project (No. 2023ZD0504600 to HW, JY, MD, and KC); Capital ’ s Funds for Health Improvement and Research (2022-1-4052 to HW, MD, JY, PZ, and KC); the National High-Level Hospital Clinical Research Funding (BJYY-2023-070 to HW, MD, JY, TW, and KC); the National Natural Science Foundation of China (No. 82170396 to HW and KC); and the CAMS Innovation Fund for Medical Sciences (2021-I2M-1-050 to HW).

We also extend our sincere gratitude to all the reviewers for their constructive feedback on our manuscript. Their insightful comments and suggestions have significantly enhanced the quality and clarity of this work.

## References

1 Khan MS, Shahid I, Bennis A, Rakisheva A, Metra M, Butler J. Global epidemiology of heart failure. Nat Rev Cardiol 2024; 21(10): 717–34.

2 Savarese G, Becher PM, Lund LH, Seferovic P, Rosano GMC, Coats AJS. Global burden of heart failure: a comprehensive and updated review of epidemiology. Cardiovasc Res 2023; 118(17): 3272–87.

3 Wang H, Chai K, Du M, et al. Prevalence and incidence of heart failure among urban patients in china: a national population-based analysis. Circ Heart Fail 2021; 14(10): e008406.

4 Hao G, Wang X, Chen Z, et al. Prevalence of heart failure and left ventricular dysfunction in china: the china hypertension survey, 2012-2015. Eur J Heart Fail 2019; 21(11): 1329–37.

5 GBD 2021 forecasting collaborators. Burden of disease scenarios for 204 countries and territories, 2022-2050: a forecasting analysis for the global burden of disease study 2021. Lancet 2024; 403(10440): 2204–56.

6 Roth GA, Mensah GA, Johnson CO, et al. Global burden of cardiovascular diseases and risk factors, 1990-2019: update from the GBD 2019 study. J Am Coll Cardiol 2020; 76(25): 2982–3021.

7 Piepoli MF, Adamo M, Barison A, et al. Preventing heart failure: a position paper of the heart failure association in collaboration with the european association of preventive cardiology. Eur J Heart Fail 2022; 24(1): 143–68.

8 Basic C, Rosengren A, Alehagen U, et al. Young patients with heart failure: clinical characteristics and outcomes. Data from the swedish heart failure, national patient, population and cause of death registers. Eur J Heart Fail 2020; 22(7): 1125–32.

9 Jamison DT, Summers LH, Chang AY, et al. Global health 2050: the path to halving premature death by mid-century. Lancet 2024; 404(10462): 1561–614.

10 Jürgens V, Ess S, Cerny T, Vounatsou P. A bayesian generalized age-period-cohort power model for cancer projections. Stat Med 2014; 33(26): 4627–36.

11 Chen S, Cao Z, Nandi A, et al. The global macroeconomic burden of alzheimer’s disease and other dementias: estimates and projections for 152 countries or territories. Lancet Glob Health 2024; 12(9): e1534–43.

12 Chen S, Kuhn M, Prettner K, et al. The global economic burden of chronic obstructive pulmonary disease for 204 countries and territories in 2020-50: a health-augmented macroeconomic modelling study. Lancet Glob Health 2023; 11(8): e1183–93.

13 Chen S, Cao Z, Prettner K, et al. Estimates and projections of the global economic cost of 29 cancers in 204 countries and territories from 2020 to 2050. JAMA Oncol 2023; 9(4): 465–72.

14 Wang H, Li Y, Chai K, et al. Mortality in patients admitted to hospital with heart failure in china: a nationwide cardiovascular association database-heart failure centre registry cohort study. Lancet Glob Health 2024; 12(4): e611–22.

15 Mcdonagh TA, Metra M, Adamo M, et al. 2021 ESC guidelines for the diagnosis and treatment of acute and chronic heart failure. Eur Heart J 2021; 42(36): 3599–726.

16 Heidenreich PA, Bozkurt B, Aguilar D, et al. 2022 AHA/ACC/HFSA guideline for the management of heart failure: a report of the american college of cardiology/american heart association joint committee on clinical practice guidelines. Circulation 2022; 145(18): e895–1032.

17 Chinese medical association, Chinese medical association publishing house, Chinese society of general practice, Editorial board of chinese journal of general practitioners of chinese medical association, Electrophysiology and cardiac function branch of chinese society of geriatrics, Expert group on the Chinese guidelines for the diagnosis and management of heart failure in primary care (2024), Chinese guideline for the diagnosis and management of heart failure in primary care (2024). Chin J Gen Pract 2024(06): 549–77

18 Ledwidge M, Gallagher J, Conlon C, et al. Natriuretic peptide-based screening and collaborative care for heart failure: the STOP-HF randomized trial. Jama 2013; 310(1): 66–74.

19 Li C, Chen K, Cornelius V, et al. Applicability and cost-effectiveness of the systolic blood pressure intervention trial (SPRINT) in the chinese population: a cost-effectiveness modeling study. PLoS Med 2021; 18(3): e1003515.

20 Wright JTJ, Williamson JD, Whelton PK, et al. A randomized trial of intensive versus standard blood-pressure control. N Engl J Med 2015; 373(22): 2103–16.

21 Zhang W, Zhang S, Deng Y, et al. Trial of intensive blood-pressure control in older patients with hypertension. N Engl J Med 2021; 385(14): 1268–79.

22 Cao TH, Tay WT, Jones DJL, et al. Heart failure with improved versus persistently reduced left ventricular ejection fraction: a comparison of the BIOSTAT-CHF (european) study with the ASIAN-HF registry. Eur J Heart Fail 2024; 26(12): 2518–28.

23 Tsao CW, Aday AW, Almarzooq ZI, et al. Heart disease and stroke statistics-2022 update: a report from the american heart association. Circulation 2022; 145(8): e153–639.

24 Bundgaard JS, Mogensen UM, Christensen S, et al. The economic burden of heart failure in denmark from 1998 to 2016. Eur J Heart Fail 2019; 21(12): 1526–31.

25 Chong B, Jayabaskaran J, Jauhari SM, et al. Global burden of cardiovascular diseases: projections from 2025 to 2050. Eur J Prev Cardiol 2024.

26 Liu J, Liu M, Chai Z, et al. Projected rapid growth in diabetes disease burden and economic burden in china: a spatio-temporal study from 2020 to 2030. Lancet Reg Health West Pac 2023; 33: 100700.

27 Tromp J, Paniagua SMA, Lau ES, et al. Age dependent associations of risk factors with heart failure: pooled population based cohort study. Bmj 2021; 372: n461.

28 Ledwidge MT, O’Connell E, Gallagher J, et al. Cost-effectiveness of natriuretic peptide-based screening and collaborative care: a report from the STOP-HF (st vincent’s screening to prevent heart failure) study. Eur J Heart Fail 2015; 17(7): 672–9.

29 Zhou Y, Li C, Wang M, et al. Universal health coverage in china: a serial national cross-sectional study of surveys from 2003 to 2018. Lancet Public Health 2022; 7(12): e1051–63.

30 Zhang H, Huo X, Ren L, et al. Design and rationale of the comprehensive intelligent hypertension management SyStem (CHESS) evaluation study: a cluster randomized controlled trial for hypertension management in primary care. Am Heart J 2024; 273: 90–101.

31 Sun Y, Mu J, Wang DW, et al. A village doctor-led multifaceted intervention for blood pressure control in rural china: an open, cluster randomised trial. Lancet 2022; 399(10339): 1964–75.

32 Greene SJ, Ayodele I, Pierce JB, et al. Eligibility and projected benefits of rapid initiation of quadruple therapy for newly diagnosed heart failure. JACC Heart Fail 2024; 12(8): 1365–77.

33 Davis JA, Booth D, Mcewan P, et al. Cost-effectiveness of dapagliflozin for patients with heart failure across the spectrum of ejection fraction: a pooled analysis of DAPA-HF and DELIVER data. Eur J Heart Fail 2024; 26(3): 664–73.

34 Sumarsono A, Xie L, Keshvani N, et al. Sex disparities in longitudinal use and intensification of guideline-directed medical therapy among patients with newly diagnosed heart failure with reduced ejection fraction. Circulation 2024; 149(7): 510–20.

35 Banka G, Heidenreich PA, Fonarow GC. Incremental cost-effectiveness of guideline-directed medical therapies for heart failure. J Am Coll Cardiol 2013; 61(13): 1440–6.

36 Tromp J, Bamadhaj S, Cleland JGF, et al. Post-discharge prognosis of patients admitted to hospital for heart failure by world region, and national level of income and income disparity (REPORT-HF): a cohort study. Lancet Glob Health 2020; 8(3): e411–22.

37 Man JP, Koole MAC, Meregalli PG, et al. Digital consults in heart failure care: a randomized controlled trial. Nat Med 2024; 30(10): 2907–13.

38 Liu H, Yin P, Qi J, Zhou M. Burden of non-communicable diseases in china and its provinces, 1990-2021: results from the global burden of disease study 2021. Chin Med J (Engl) 2024; 137(19): 2325–33.

