## Supplementary material for "The Burden and Impact of Intervention Strategies for Heart Failure in China (2025–2035): A Health-Augmented Macroeconomic Modeling Study"

**A：Modeling details I**

The construction of this model is based on the health-augmented macroeconomic modeling framework used in the study titled *"The Global Economic Burden of Chronic Obstructive Pulmonary Disease for 204 Countries and Territories, 2020–50: A Health-Augmented Macroeconomic Modelling Study."* [1]This framework has been extensively applied in various studies, including assessments of the economic burden of chronic diseases in China, Japan, South Korea, the United States, and European countries;[2] analyses of the burden of road injuries;[3] investigations into the burden of cancer;[4] and evaluations of the economic impacts of risk factors such as tobacco use and air pollution.[5,6]

This study aims to quantify the impact of heart failure (HF) on economic output through two primary pathways: the direct economic burden of healthcare expenditures and productivity losses attributable to mortality and morbidity. To this end, we conducted the following analyses:

**Step 1. Quantification of HF Disease Burden and Macroeconomic Pathways**

We systematically quantified the burden of HF through mortality rates, prevalence estimates, and direct healthcare expenditures. Concurrently, the pathways by which HF impacts macroeconomic output were identified, encompassing increased healthcare resource allocation, workforce productivity losses from morbidity and premature mortality, and long-term constraints on capital accumulation due to expenditure reallocation. This multi-dimensional assessment established the foundation for modeling HF's economic repercussions.

**Step 2. Economic Projection Framework**

We developed two distinct economic scenarios using a macroeconomic production function:

(i) **Status quo scenario**: Projected aggregate output $Y_t$ under existing conditions without targeted interventions to reduce HF-related mortality or prevalence in year $t$.

(ii) **Counterfactual scenario**: Hypothetical aggregate output $\overline{Y_t}$ assuming complete elimination of HF at zero cost in year $t$.

The projections were stratified into two components:

a) **Effective labor supply dynamics,** accounting for workforce productivity impacts due to HF morbidity and mortality.

b) **Physical capital accumulation,** reflecting investment trajectories influenced by healthcare expenditure reallocations.

**Step 3. Economic Impact Analysis**

For each year, we computed the annual gross domestic product (GDP) differential between the two scenarios. The total discounted economic loss was derived as the cumulative GDP differential over the study period, discounted to present value using a standard rate to reflect temporal economic trade-offs.

**Production Function**

We consider a discrete-time economy ( $t = 1,2,\ldots,\infty$ ) with an aggregate production function derived from Lucas (1988)[7]:

$$Y_t = A_t K_t^{\alpha} H_t^{1-\alpha}, \quad (1)$$

where $Y_t$ denotes aggregate output; $A_t$ represents the technological level at time $t$, which is assumed to evolve exogenously; $K_t$ is the physical capital stock; and $H_t$ captures aggregate human capital. The parameter $\alpha$ reflects the elasticity of final output with respect to physical capital. The Solow growth model, foundational to the World Health Organization's (WHO) original EPIC (Economic Projection Integrated with Health) macroeconomic framework, restricts its analytical scope to physical capital and unadjusted labor inputs as primary determinants of economic output.[8,9] In contrast, the aggregate production function explicitly acknowledges that economic output is generated not merely through these conventional factors but also via health-augmented labor inputs. Health status, as a critical dimension of human capital, is thereby integrated as an essential determinant of productive efficiency within this expanded paradigm.

**Step 1. Quantification of HF Disease Burden and Macroeconomic Pathways**

The disease burden of HF was systematically quantified through three dimensions: (i) mortality rates, (ii) prevalence estimates, and (iii) direct healthcare expenditures (e.g., hospitalization, pharmacotherapy). Concurrently, the macroeconomic pathways linking HF to GDP reduction were formalized, integrating both physical capital constraints and effective labor supply dynamics into the analytical framework.

HF-related treatment costs ($TC_t$) directly reduce national savings available for capital accumulation.

Physical capital evolves according to the following dynamic equation:

$$K_{t+1} = (1-\delta)K_t + Y_t - C_t - TC_t = (1-\delta)K_t + s_t Y_t, \quad (2)$$

where $\delta$ demotes the depreciation rate, $s_t$ refers to the saving rate, $TC_t$ represents the costs of ongoing treatment of HF, and $C_t$ is aggregate consumption. From Equation (2), it follows that the saving rate is defined as

$$s_t = 1 - \frac{C_t + TC_t}{Y_t}.$$

Aggregate output $Y_t$ is allocated to three purposes: (i) consumption ($C_t$), (ii) savings (for capital accumulation), and (iii) HF treatment expenditures. Elevated $TC_t$ reduces $s_t$ , thereby diminishing the reinvestment into physical capital ($K_{t+1}$) and constraining long-term economic growth.

Mortality and prevalence reduce workforce productivity through two distinct channels: mortality-driven population decline and morbidity-driven labor participation decline.

Aggregate human capital $H_t$ in the production function (1) is then defined as the sum over the age- and gender-specific effective labor supply:

$$H_t = \sum_{i=0}^{1} \sum_{a=15}^{R_i} h_t^{(a,i)} l_t^{(a,i)} N_t^{(a,i)}, \quad (3)$$

where $h_t^{(a,i)}$, $l_t^{(a,i)}$ and $N_t^{(a,i)}$ represent individual human capital endowment, labor participation, and population, respectively. Here $a$ represents the age group, and $i$ indicates gender ( $i$=0 for male, $i$=1 for female). This sum is calculated over the age-specific effective labor supply of each age group from age 15 up to their retirement at age $R$, i.e., for $R \in [15, R]$. Children younger than 15 and retirees older than do not work. Specifically, in this model, based on China's actual retirement conditions, the retirement age for females is set to 55 years ( $R_1 = 55$), and for males, it is set to 60 years ( $R_0 = 60$).

In Equation (3), $N_t^{(a,i)}$ can be calculated using the following formula:

$$N_t^{(a,i)} = [1 - \sigma_{t-1}^{(a-1,i)}]N_{t-1}^{(a-1,i)}, \tag{4}$$

where $\sigma_t^{(a,i)}$ is the overall mortality rate of age and sex group $(a, i)$ at time ˆ. Let $\sigma_{r,t}^{(a,i)}$ denote the mortality rate of people in age and sex group R$(a, i)$ due to HF and let $\sigma_{-r,t}^{(a,i)}$ be the overall mortality rate due to causes other than HF. Then, we have the following relationship:

$$(1 - \sigma_t^{(a,i)}) = (1 - \sigma_{r,t}^{(a,i)})(1 - \sigma_{-r,t}^{(a,i)}). \tag{5}$$

This can clearly show that the mortality effect of HF is to reduce the population $N_t^{(a,i)}$.

The reduction of the labor participation rate $l_t^{(a,i)}$ captures the prevalence effect because people with an illness typically reduce their labor supply. Based on clinical experience, the model assumes that individuals with HF are unable to participate in the labor force from the year of onset until they are either cured or they die.

**Step 2. Economic Projection Framework**

We developed two distinct economic scenarios to quantify the macroeconomic impact of HF:

(i) Status quo scenario: Aggregate output $Y_t$ reflects current conditions without targeted interventions to reduce HF mortality or prevalence. Projections for $Y_t$ were sourced from the International Monetary Fund's World Economic Outlook Database.[10]

(ii) Counterfactual scenario: Hypothetical aggregate output $\overline{Y_t}$ assumes complete HF elimination at zero cost in year $t$. This scenario is modeled as:

$$\overline{Y_t} = A_t \overline{K_t}^{\alpha} \overline{H_t}^{1-\alpha}, \tag{6}$$

where $\overline{K_t}$ and $\overline{H_t}$ denote the physical capital stock and aggregate human capital under HF elimination, respectively. Technological progress $A_t$ is assumed identical across both scenarios and derived from the status quo output:

$$A_t = \frac{Y_t}{K_t^{\alpha} H_t^{1-\alpha}}.$$

Here, $K_t$ and $H_t$ are calculated via Equations (2) and (3), respectively (see Step 1). Regarding $h_t^{(a,i)}$ in (3), we followed Mincer (1974)[11] and constructed the average human capital of the cohort aged $a$ according to an exponential function of education and work experience:

$$h_t^{(a,i)} = exp[\eta_1(ys_t^{(a,i)}) + \eta_2(a - ys_t^{(a,i)} - 5) + \eta_3(a - ys_t^{(a,i)} - 5)^2], \tag{7}$$

where $\eta_1$ is the semi-elasticity of human capital with respect to average years of education as given by $ys_t^{(a,i)}$, and $\eta_2$ and $\eta_3$ are the semi-elasticities of human capital with respect to experience of the workforce $(a - ys_t^{(a,i)} - 5)$ and experience of the workforce squared $(a - ys_t^{(a,i)} - 5)^2$, respectively. Here, we assumed a school entry age of 5 years throughout. $ys_t^{(a,i)}$ is the average years of education of people in age and sex group $(a, i)$ in year *t*.

Capital accumulation under HF elimination ($\overline{K_t}$) follows:

$$\overline{K_{t+1}} = \overline{s_t}\,\overline{Y_t} + (1-\delta)\,\overline{K_t}, \tag{8}$$

with the saving rate adjusted to incorporate reallocated HF treatment costs:

$$\overline{s_t}\,\overline{Y_t} = \overline{I} = \overline{Y_t} - \overline{C_t} = s_t\,\overline{Y_t} + \chi TC_t. \tag{9}$$

Here, $\chi$ represents the fraction of the treatment cost ($TC_t$) diverted to savings.

By eliminating HF, resources previously allocated to treatment ($TC_t$) are redistributed between savings and consumption. For tractability, we assume: A fixed baseline saving rate $s_t$ and additional savings from $\chi TC_t$, reflecting the opportunity cost of disease mitigation.

This framework aligns with methodologies from Bloom et al. (2020)[2] and Chen et al. (2018)[12], explicitly modeling the income effect of healthcare expenditure reallocation.

Total annual treatment costs for HF ($TC_t$) are calculated as:

$$TC_t = \sum_{i=0}^{1}\sum_{a=15}^{100} tc_t^{(a,i)} \varphi_t^{(a,i)} N_t^{(a,i)}, \tag{10}$$

where $tc_t^{(a,i)}$, $\varphi_t^{(a,i)}$ and $N_t^{(a,i)}$ represent the HF treatment costs per capita, the prevalence rate of HF and the population in age and sex group $(a, i)$ in year $t$, respectively.

For $\overline{H_t}$, similar to equation (3), we have:

$$\overline{H}_t = \sum_{i=0}^{1}\sum_{a=15}^{R_i} h_t^{(a,i)}\,\overline{l_t^{(a,i)}}\,\overline{N_t^{(a,i)}}, \tag{11}$$

where $h_t^{(a,i)}$ in both the counterfactual and status quo scenarios is similar, therefore it is calculated via (2).

From equations (4) and (5), we can obtain:

$$N_t^{(a,i)} = [1-\sigma_{-r,t-1}^{(a-1,i)}][1-\sigma_{r,t}^{(a,i)}]N_{t-1}^{(a-1,i)}.$$

In the counterfactual scenario, $\sigma_{r,t}^{(a,i)}$ is assumed to be zero, so the calculation for $\overline{N_t^{(a,i)}}$ is as follows:

$$\overline{N_t^{(a,i)}} = [1-\sigma_{-r,t-1}^{(a-1,i)}]\overline{N_{t-1}^{(a-1,i)}},\quad \overline{N_{2025}^{(a,i)}} = N_{2025}^{(a,i)},\quad \overline{N_t^{(0,i)}} = N_t^{(0,i)}.$$

Following Bloom et al. (2020)[2], for $t$>2025, the loss of labor due to mortality accumulates over the years according to

$$\overline{N_t^{(a,i)}} = N_t^{(a,i)} \Big/ \prod_{\tau=0}^{\min\{t-2025,a\}-1}[1-\sigma_{r,t-1-\tau}^{(a-1-\tau,i)}].$$

Since we cannot directly obtain the mortality rate from HF, we have broken it down into the product of the case fatality rate of HF and the prevalence of HF:

$$\sigma_{r,t}^{(a,i)} = \omega_t^{(a,i)}\varphi_t^{(a,i)}, \tag{12}$$

where $\omega_t^{(a,i)}$ and $\varphi_t^{(a,i)}$ represent the case fatality rate (proportion of HF cases resulting in death) and the prevalence rate of HF in age and sex group $(a,i)$ in year $t$.

In equation (11), $\overline{l_t^{(a,i)}}$ represents the labor participation rate in the counterfactual scenario. In the status quo scenario, part of the labor force is excluded from the labor market due to illness, which reduces the labor participation rate. Assuming the prevalence rate in the labor force is equal to the prevalence rate in the general population, and that all individuals with HF no longer provide labor, we can obtain the relationship between $l_t^{(a,i)}$ and $\overline{l_t^{(a,i)}}$ as follows:

$$l_t^{(a,i)} = (1-\varphi_t^{(a,i)})\overline{l_t^{(a,i)}}. \tag{13}$$

This adjustment reflects the reintegration of previously ill individuals into the labor force under HF elimination. Thus, we obtain:

$$\overline{l_t^{(a,i)}} = l_t^{(a,i)} \Big/ (1-\varphi_t^{(a,i)}).$$

**Step 3. Economic Impact Analysis**

The annual gross domestic product (GDP) differential between the status quo and counterfactual scenarios was computed for each year $t$. To quantify the long-term economic burden of HF, the total discounted economic loss ($\Delta Y$) was derived as the cumulative difference in projected GDP, adjusted for temporal valuation using a standard discount rate:

$$\Delta Y = \sum_{t=2025}^{2035}\left[\left(\overline{Y_t} - Y_t\right) \Big/ (1+discount\ rate)^{t-2025}\right]$$

**B: Modeling details II**

To estimate the economic burden reduction attributable to targeted interventions for HF, we adapted methodological approaches from Bloom et al. (2020)[2]. Two economic scenarios were constructed:

(i) Status quo scenario: Aggregate output $\tilde{Y}_t$ reflects baseline conditions without additional interventions to mitigate HF-related mortality or morbidity.

(ii) Counterfactual scenario: Aggregate output $\overline{Y_t}$ assumes implementation of a specific intervention in year $t$, modeled as:

$$\overline{Y_t} = A_t \overline{K_t}^{\alpha} \overline{H_t}^{1-\alpha},$$

where $\overline{K_t}$ and $\overline{H_t}$ denote physical capital stock and health-augmented human capital under the intervention, respectively.

(1) **Intensive Blood Pressure (BP) Control**

First, we calculate the effect of this intervention on the reduction of HF prevalence rate.

Here, we note that the intervention leads to a reduction in morbidity from $M_{before}$ to $M_{after}$, resulting in a reduction in the number of newly diagnosed HF patients in year $t$, denoted as $Reduced_t$. We have:

$$Reduced_t = (M_{before} - M_{after})TP_t \times Intervene\_cover,$$

where $TP_t$ represents the target population in year $t$ (the number of hypertensive patients in year $t$), $Intervene\_cover$ represents the intervention coverage.

When calculating the impact on the prevalence rate of HF, we need the average duration of HF. According to the China Cardiovascular Association (CCA) Database-Heart Failure Centre Registry [13], the average duration of HF is 3.914 years. For simplicity, the model assumes a duration of 4 years. Thus, the impact of disease reduction is calculated as:

$$\rho_t = \frac{\sum_{\tau=0}^{min\{t-2025,3\}} Reduced_{t-\tau}}{Total\_patients_t}, \quad (14)$$

where $\rho_t$ represents the relative reduction in the prevalence rate in year $t$, $\sum_{\tau=0}^{t=min\{t-2025,3\}} Reduced_{t-\tau}$ equals to the reduction in the number of HF patients in year $t$ and

$$Total\_patients_t = \sum_{i=0}^{1} \sum_{a=15}^{100} \varphi_t^{(a,i)} N_t^{(a,i)} \quad (15)$$

represents the total number of patients in year $t$ in the status quo scenario without any additional interventions. Here $\varphi_t^{(a,i)}$ and $N_t^{(a,i)}$ correspond to the prevalence rate and population for each age and gender group in the status quo scenario.

Next, we calculate the impact of this intervention on effective labor supply. Equation (12) and (13) are modified as follows:

$$\sigma_{r,t}^{(a,i)} = \omega_t^{(a,i)} \rho_t \varphi_t^{(a,i)},$$

$$l_t^{(a,i)} = (1 - \rho_t \varphi_t^{(a,i)}) \overline{l_t^{(a,i)}},$$

where $\rho\varphi_t^{(a,i)}$ represents the reduction in disease rates due to the intervention in year $t$, $\omega_t^{(a,i)}$ represents the case fatality rate and $l_t^{(a,i)}$ represents the labor participation for each age and gender group $(a, i)$.

Finally, we calculate the impact of this intervention on physical capital accumulation. Equation (9) is modified as follows:

$$\overline{s_t}\,\overline{Y_t} = \overline{I} = \overline{Y_t} - \overline{C_t} = s_t \overline{Y_t} + \chi\ (\rho_t TC_t - IC_t),$$

where $\rho_t TC_t$ represents the reduction of HF treatment costs in year $t$ due to the decline in prevalence, while $IC_t$ is the cost of intervention in year $t$,calculated as:

$$IC_t = TP_t \times Intervene\_cover \times ic_t,$$

where $ic_t$ is the cost of intervention per person in year $t$.

Other equations remain unchanged. Thus, we can use the previous macroeconomic model to calculate the economic burden reduction due to this intervention.

Specifically, for the Intensive BP Control intervention, we assume that the target population remains constant at $TP_t$=245 million, and the cost of intervention per person per year does not change.

(2) **BNP Screening**

It is similar to the calculated BP intervention.

The target population for this intervention includes all individuals aged 40 years and older. Specifically, the target population is calculated as:

$$TP_t = \sum_{i=0}^{1} \sum_{a=40}^{100} N_t^{(a,i)}.$$

This intervention specifically targets individuals over 40, requiring adjustments to age-related metrics starting from 40. The following adjustments are made:

We adjust equation (15) to calculate the total number of patients as:

$$Total\_patients_t = \sum_{i=0}^{1} \sum_{a=40}^{100} \varphi_t^{(a,i)} N_t^{(a,i)}.$$

The intervention affects the effective labor supply for individuals over 40, leading to the adjustment of equation (11) to:

$$\overline{H_t} = \sum_{i=0}^{1} \sum_{a=40}^{R_i} h_t^{(a,i)} \overline{l_t^{(a,i)}}\, \overline{N_t^{(a,i)}}.$$

The treatment costs for individuals over 40 are influenced by the intervention, resulting in the adjustment of equation (10) to:

$$TC_t = \sum_{i=0}^{1} \sum_{a=40}^{100} tc_t^{(a,i)} \varphi_t^{(a,i)} N_t^{(a,i)}.$$

All other aspects of the calculation remain identical to those of Intensive Blood Pressure Control. This structured approach ensures that the BNP screening intervention is accurately modeled and its impact on the specified population is effectively assessed.

(3) **Guideline-Directed Medical Therapy (GDMT) for HF with reduced ejection fraction (HFrEF)**

The target population for this intervention consists of newly diagnosed HFrEF patients, for whom we aim to increase GDMT coverage.

$\Delta Intervene\_cover$ is defined as:

$$\Delta Intervene\_cover = Intervene\_cover_{target} - Intervene\_cover_{before}.$$

In the status quo scenario, we assume that the proportion of newly diagnosed HFrEF patients receiving GDMT is $Intervene\_cover_{before}$, while in the counterfactual scenario, the proportion of newly diagnosed HFrEF patients receiving GDMT will increase to $Intervene\_cover_{target}$. Consequently, the increase in GDMT coverage is modeled using $\Delta Intervene\_cover$, rather than assuming it is $Intervene\_cover_{target}$.

For modeling convenience, we assume that all patients receiving GDMT have two possible outcomes:1. A proportion of patients, determined by the $Recovery\_rate$, recover within one year after treatment. These patients can return to the labor market but continue receiving GDMT until 2035, with corresponding annual costs. 2. The remaining patients will continue to receive GDMT throughout the progression of the disease, though they will not achieve complete remission (assuming the disease progression lasts approximately four years).

Based on the assumptions above, we calculate the economic burden reduction due to this intervention as follows:

The total number of patients in year $t$ in the status quo scenario is expressed as:

$$Total\_patients_t = \sum_{i=0}^{1} \sum_{a=15}^{100} \varphi_t^{(a,i)} N_t^{(a,i)},$$

where $\varphi_t^{(a,i)}$ and $N_t^{(a,i)}$ correspond to the prevalence rate of HF and population for each age and gender group $(a,i)$. Thus $Total\_patients_t$ represents the total number of HF patients in year $t$ in the status quo scenario. For this intervention, the target population is newly diagnosed HFrEF patients. The target population, denoted as $TP_t$, is calculated as:

$$TP_t = \frac{HFrEF\_proportion \times Total\_patients_t}{the\ average\ duration\ of\ heart\ failure},$$

where $\frac{Total\_patients_t}{the\ average\ duration\ of\ heart\ failure}$ is the number of newly diagnosed HF patients in year $t$,

$HFrEF\_proportion$ represents the proportion of HFrEF patients among all HF patients.
We then calculate the number of patients newly receiving GDMT annually in the counterfactual scenario, denoted as $Patient\_inter_t$ , which is:

$$Patient\_inter_t = \Delta Intervene\_cover \times TP_t.$$

From this, we can calculate the number of newly cured patients in year $t+1$ due to the intervention, denoted as $Reduced_{t+1}$ , which is calculated as:

$$Reduced_{t+1} = Patient\_inter_t \times Recovery\_rate,$$

where $Recovery\_rate$ represents the proportion of patients receiving GDMT who recover within one year.
Thus, the impact of disease reduction is calculated as:

$$\rho_t = \frac{\sum_{\tau=0}^{min\{t-2025,3\}} Reduced_{t-\tau}}{Total\_patients_t},$$

where $\rho_t$ represents the relative reduction in the prevalence rate in year $t$. As can be seen, this is consistent with equation (14). Then, we calculate the impact of this intervention on effective labor supply, which follows the same approach as the calculation of Intensive Blood Pressure Control.
Then we calculate the impact of this intervention on physical capital accumulation. Equation (9) is modified as follows:

$$\overline{s_t}\,\overline{Y_t} = \overline{I} = \overline{Y_t} - \overline{C_t} = s_t Y_t + \chi\ (\rho_t TC_t - IC_t),$$

where $\rho_t TC_t$ represents the reduction of HF treatment costs in year $t$ due to the decline in prevalence, while $IC_t$ is the cost of intervention in year $t$, calculated as:

$$IC_t = \left(\sum_{\tau=2026}^{t} Reduced_\tau + \sum_{\tau=0}^{min\{t-2025,3\}} (Patient\_inter_{t-\tau} - Reduced_\tau)\right) \times ic_t,$$

where $ic_t$ is the cost of intervention per person in year $t$. The reason for this calculation is that the number of patients who need to receive GDMT each year is composed of two parts. (i) Patients who have received intervention and recovered within one year will continue to receive GDMT, and the number of patients in this part is $\sum_{\tau=2026}^{t} Reduced_\tau$ in year $t$. (ii) Patients who have received the intervention but are not cured within one year, they will receive GDMT throughout the course of the disease, and the number of patients in this segment is $\sum_{\tau=0}^{min\{t-2025,3\}} (Patient\_inter_{t-\tau} - Reduced_\tau)$ in year $t$。

The rest of the calculation is the same as for Intensive BP Control.

**C: Decomposing the Economic Burden: Contributions of Physical Capital Accumulation and Effective Labor Supply Dynamics**

From equation (1):

$$Y_t = A_t K_t^{\alpha} H_t^{1-\alpha}$$

the changes in output over time can be expressed as:

$$\frac{\Delta Y_t}{Y_t} = \frac{\Delta A_t}{A_t} + \alpha \frac{\Delta K_t}{K_t} + (1-\alpha)\frac{\Delta H_t}{H_t}.$$

In this model, we assume that $A_t$ evolves exogenously ($\Delta A_t = 0$), then we have:

$$\overline{Y_t} - Y_t = \alpha \frac{\overline{K_t} - K_t}{K_t} Y_t + (1-\alpha)\frac{\overline{H_t} - H_t}{H_t} Y_t.$$

Thus, we can break down the change in $Y_t$ into two components: one driven by the change in physical capital ($\alpha \frac{\overline{K_t} - K_t}{K_t} Y_t$) and the other by the change in labor supply $((1-\alpha)\frac{\overline{H_t} - H_t}{H_t} Y_t)$.

The contribution of physical capital to the economic burden is given by:

$$Proprotion_{K_t} = \frac{\alpha \frac{\overline{K_t} - K_t}{K_t} Y_t}{\overline{Y_t} - Y_t}.$$

Similarly, the contribution of labor supply is:

$$Proprotion_{H_t} = \frac{(1-\alpha)\frac{\overline{H_t} - H_t}{H_t} Y_t}{\overline{Y_t} - Y_t}.$$

We can also compute the cumulative contributions over the period from 2025 to 2035:

$$Total\_Proprotion_{K_t} = \sum_{t=2025}^{2035}\left[\left(\frac{Contribution_{K_t}}{\overline{Y_t} - Y_t}\right) \div (1 + discount\ rate)^{t-2025}\right],$$

$$Total\_Proprotion_{H_t} = \sum_{t=2025}^{2035}\left[\left(\frac{Contribution_{H_t}}{\overline{Y_t} - Y_t}\right) \div (1 + discount\ rate)^{t-2025}\right].$$

**D：Data description**

**Education**

Age- and gender- specific educational attainment data were sourced from the Barro-Lee Educational Attainment Database, which provides data in five-year age groups up to 2010.[14] However, no age- and gender- specific data are available for the period from 2010 to 2035.

Missing observations are filled using forward and backward extrapolation of census and survey data on educational attainment, following the Barro-Lee Database methodology.[14] It is assumed that an individual's educational attainment remains constant between the ages of 25 and 64, and that mortality rates are uniform across all individuals, regardless of educational attainment. Consequently, for age groups between 25 and 64, missing attainment data are filled by using the attainment data from the younger age group of the previous period (forward) as a benchmark, or from the older age group of the succeeding period (backward). We adopted this approach for our analysis. For age groups between 30 and 64, we directly used the attainment data from the younger age group of the previous period to fill the gaps for the years 2010–2035. For the 25–30 age group, we assumed that individuals in this group might still pursue higher education, but that attainment levels for primary and secondary education would remain unchanged. As a result, we used the attainment data from the younger age group of the previous period to fill the gaps for primary and secondary education for the 2010–2035 period.

For the university education levels of the two youngest age groups (15–19 and 20–24), as well as for the 25–30 age group, we referred to the assumptions made by Simiao Chen (2023). Specifically, we projected educational attainment by assuming the same growth rate for each age and sex group as observed in the 2000–2010 period.

For age groups above 65, no predictions were made for educational attainment, as this data was not required in the model. This omission aligns with the Chinese pension system, which mandates retirement and excludes individuals aged 65 or older from formal employment, thereby rendering educational attainment irrelevant to labor force participation in this demographic.

Since the Barro-Lee database presents data in five-year intervals, linear interpolation was employed to extend the estimates for each year.

**Prevalence**

Projected HF Prevalence in China was derived from the Global Burden of Disease (GBD) Study 2021, adjusted for China-specific demographic and risk factor trends.[15] Detailed methodological specifications regarding data integration, modeling assumptions, and sensitivity analyses are provided in the Methods section of the main text.

**Case fatality rate**

Age- and gender-specific case fatality rates were calculated based on the China Cardiovascular Association (CCA) Database-Heart Failure Centre Registry.[13] We assume that the case fatality rates from 2025 to 2035 will remain unchanged and be consistent with the levels observed in 2021.

**GDP projection**

The GDP estimates (in constant 2017 international dollars or INT$) from 2025 to 2035 were from International Monetary Fund World Economic Outlook Database.[10]

**Physical capital**

The physical capital stock (in 2017 INT$) was sourced from the Penn World Table version 10.01.[16] However, since the data in this database are only updated through 2019, we utilized GDP data (in constant 2017 international dollars or INT$) from the International Monetary Fund World Economic Outlook Database for the period 2019 to 2024.[10] For the years 2025 to 2035, we used GDP estimates to derive the physical capital stock for this period using equation (2). The value for the output elasticity of physical capital (the percentage change in output for a 1% change in the physical capital stock) following standard economic estimates is also from the Penn World Table version 10.01.[16]

**Labor participation rate**

The labor participation rates were obtained from the International Labour Organization (ILO) database (Labor Force Participation Rate by Sex and Age-ILO Modeled Estimates, Nov. 2024 (%)).[17] Since this database only provides labor participation rate projections up to 2026, we assumed that the growth rate of the labor participation rate beyond 2026 would remain consistent with the growth observed during the period from 2016 to 2026.

**Population projection**

The population estimates were obtained from 2024 Revision of World Population Prospects prepared by the Population Division of the Department of Economic and Social Affairs of the United Nations Secretariat.[18]

**Saving rate**

The saving rate was obtained from the Worldbank database.[19] In the model, we assume that the saving rate from 2025 to 2035 remains unchanged and is equal to the average saving rate observed during the period from 2014 to 2023.

**Treatment costs**

We utilized data from the National Urban Employee Basic Medical Insurance program across six provinces in China (2017) to calculate the per capita HF treatment costs by age group and gender for 2017 (converted to 2017 INT$).[20] These figures were then used as the national per capita HF treatment costs by age group and gender. Based on a more conservative estimate, we assume that the per capita HF treatment costs from 2025 to 2035 will remain unchanged and consistent with the 2017 levels.

**Discount rate**

In the main analysis, we used a discount rate of 3%. As noted in the model explanation referenced from Simiao Chen, a 3% discount rate is somewhat standard in global health and is recommended, for example, by the Panels on Cost-effectiveness in Health and Medicine.[21] Additionally, in the sensitivity analysis, we provided results for alternative discount rates of 0%, 1%, 2%, 4%, and 5%.

**E: Table S1 Parameter values and data sources in Modeling**

| Parameter | Definition | Value | Source |
|---|---|---|---|
| $\alpha$ | Capital share | 0.413747 | Penn World Table version 10.01[16] |
| $\delta$ | Depreciation rate | 0.05 | Grossmann et al. (2013)[22] |
| $\eta_1$ | Mincer elasticity of education | 0.091 | Psacharopoulos and Patrinos (2018)[23] |
| $\eta_2$ | First-degree Mincer elasticity of experience | 0.1301 | Heckman et al. (2006)[24] |
| $\eta_3$ | Second-degree Mincer elasticity of experience | -0.0023 | Heckman et al. (2006)[24] |
| $\chi$ | Fraction of treatment cost financed out of saving | Set as saving rate | Worldbank databank[19] |

**F: Table S2 Prevalence and age-standardized rates of prevalence from 1990 to 2035 in China**

| Year | Prevalence | |
|---|---|---|
| | Episode counts (95% UI) | ASR per 100 000 (95% UI) |
| 1990 | 4683340.1 (4451173.6-4915506.61) | 644.64 (623.5-665.78) |
| 1991 | 4842349.33 (4607574.62-5077124.03) | 646.75 (625.72-667.77) |
| 1992 | 5000618.54 (4763754.57-5237482.51) | 648.96 (628.04-669.88) |
| 1993 | 5156425.2 (4917865.08-5394985.31) | 651.17 (630.35-671.99) |
| 1994 | 5311676.88 (5071756.22-5551597.54) | 653.35 (632.63-674.07) |
| 1995 | 5466718.04 (5225628.81-5707807.27) | 655.43 (634.81-676.06) |
| 1996 | 5637590.12 (5395467.13-5879713.1) | 659.06 (638.51-679.61) |
| 1997 | 5831814.45 (5588716.26-6074912.64) | 664.75 (644.27-685.23) |
| 1998 | 6037006.32 (5793050.27-6280962.37) | 670.75 (650.33-691.16) |
| 1999 | 6243347.94 (5998747.44-6487948.44) | 675.34 (655.01-695.67) |
| 2000 | 6427992.39 (6182991.46-6672993.33) | 676.73 (656.5-696.97) |
| 2001 | 6595118.13 (6350775.08-6839461.18) | 674.39 (654.33-694.44) |
| 2002 | 6762610.72 (6520024.07-7005197.38) | 669.97 (650.19-689.76) |
| 2003 | 6927859.92 (6687204.64-7168515.21) | 664.95 (645.44-684.46) |
| 2004 | 7095734.51 (6856457.83-7335011.2) | 660.81 (641.53-680.09) |
| 2005 | 7277662.31 (7038531.53-7516793.09) | 659.09 (639.95-678.23) |
| 2006 | 7512842.54 (7272768.07-7752917.02) | 661.16 (642.1-680.23) |
| 2007 | 7808825.6 (7567400.49-8050250.7) | 666.23 (647.22-685.24) |
| 2008 | 8129895.2 (7886960.05-8372830.35) | 672.68 (653.71-691.64) |
| 2009 | 8467820.28 (8223515.16-8712125.4) | 678.85 (659.93-697.77) |
| 2010 | 8780269.89 (8534911.51-9025628.26) | 683.09 (664.22-701.96) |
| 2011 | 9104321.82 (8858334.05-9350309.58) | 685.77 (666.96-704.58) |
| 2012 | 9452696.65 (9206086.8-9699306.49) | 688.2 (669.46-706.95) |
| 2013 | 9810874.03 (9563557.96-10058190.1) | 690.43 (671.74-709.11) |
| 2014 | 10221745.55 (9973745.75-10469745.35) | 692.32 (673.7-710.94) |
| 2015 | 10600278.11 (10351363.08-10849193.14) | 693.51 (674.94-712.08) |
| 2016 | 11003643.36 (10753666.2-11253620.52) | 693.38 (674.87-711.89) |
| 2017 | 11384647.5 (11133636.18-11635658.83) | 692.26 (673.8-710.71) |
| 2018 | 11756864.25 (11505007.62-12008720.89) | 691.28 (672.87-709.68) |
| 2019 | 12202997.33 (11950546.62-12455448.05) | 691.58 (673.22-709.94) |
| 2020 | 12652815.55 (12400006.62-12905624.48) | 693.72 (675.4-712.04) |
| 2021 | 13099760.71 (12846880.48-13352640.93) | 693.47 (675.18-711.75) |
| 2022 | 13340654.14 (11614937.55-15066370.73) | 689.97 (597.57-782.36) |
| 2023 | 13863156.61 (11954315.21-15771998.01) | 691.53 (592.49-790.57) |
| 2024 | 14362055.7 (12128784.59-16595326.81) | 693.63 (579.25-808.01) |
| 2025 | 14857319.19 (12078735-17635903.37) | 696.32 (552.55-840.09) |
| 2026 | 15361301.8 (11836223.9-18903846.02) | 699.66 (514.79-886.15) |
| 2027 | 15957373.83 (11714188.08-20413700.86) | 703.71 (487.33-939.55) |
| 2028 | 16700218.43 (11676165.28-22160627.57) | 708.4 (461.73-994.15) |
| 2029 | 17452979.79 (11579456.9-23948626.01) | 713.68 (436.37-1046.92) |
| 2030 | 18189522.33 (11425318.63-25741607.43) | 719.62 (413.44-1097.26) |

| | | |
|---|---|---|
| 2031 | 18933372.85 (11188698.1-27582545.45) | 726.28 (390.22-1145.75) |
| 2032 | 19743994.42 (10892457.68-29575543.32) | 733.74 (365.96-1193.41) |
| 2033 | 20706121.64 (10563818.31-31871496.94) | 741.97 (340.29-1241.18) |
| 2034 | 21708489.89 (10113045.31-34345082.34) | 750.91 (312.82-1289.98) |
| 2035 | 22696489.27 (9496048.45-36936124.13) | 760.65 (283.21-1340.77) |

ASR=Annual Standardized Prevalence Ratio；UI=Uncertainty intervals

Data were derived from the Global Burden of Disease (GBD) Study 2021- adjusted for China-specific demographic and risk factor trends.

**G: Table S3 Prevalence of Heart Failure in Males in China (year 2021–2035, %)**

| Age | 2021 | 2025 | 2026 | 2027 | 2028 | 2029 | 2030 | 2031 | 2032 | 2033 | 2034 | 2035 |
|---|---|---|---|---|---|---|---|---|---|---|---|---|
| 15-19 | 0.108 (0.107-0.109) | 0.105 (0.089-0.122) | 0.105 (0.087-0.125) | 0.105 (0.085-0.128) | 0.104 (0.082-0.131) | 0.104 (0.079-0.136) | 0.104 (0.076-0.141) | 0.105 (0.073-0.146) | 0.105 (0.070-0.152) | 0.105 (0.067-0.159) | 0.106 (0.064-0.167) | 0.107 (0.061-0.175) |
| 20-24 | 0.104 (0.103-0.105) | 0.104 (0.089-0.121) | 0.104 (0.087-0.124) | 0.104 (0.084-0.127) | 0.104 (0.082-0.131) | 0.104 (0.079-0.135) | 0.104 (0.076-0.140) | 0.104 (0.072-0.145) | 0.104 (0.069-0.151) | 0.104 (0.066-0.158) | 0.105 (0.063-0.165) | 0.105 (0.060-0.172) |
| 25-29 | 0.100 (0.099-0.101) | 0.103 (0.088-0.120) | 0.103 (0.086-0.123) | 0.103 (0.084-0.126) | 0.103 (0.081-0.130) | 0.103 (0.078-0.134) | 0.104 (0.075-0.139) | 0.104 (0.072-0.145) | 0.104 (0.069-0.151) | 0.104 (0.066-0.157) | 0.105 (0.063-0.164) | 0.105 (0.060-0.172) |
| 30-34 | 0.103 (0.103-0.104) | 0.107 (0.091-0.125) | 0.108 (0.090-0.128) | 0.108 (0.087-0.132) | 0.108 (0.085-0.136) | 0.109 (0.082-0.141) | 0.109 (0.079-0.147) | 0.109 (0.076-0.153) | 0.110 (0.073-0.159) | 0.110 (0.070-0.166) | 0.111 (0.067-0.174) | 0.111 (0.063-0.183) |
| 35-39 | 0.132 (0.131-0.133) | 0.137 (0.117-0.159) | 0.137 (0.115-0.163) | 0.138 (0.112-0.169) | 0.139 (0.109-0.175) | 0.140 (0.106-0.181) | 0.140 (0.102-0.189) | 0.141 (0.098-0.197) | 0.142 (0.094-0.206) | 0.143 (0.090-0.215) | 0.144 (0.086-0.226) | 0.145 (0.082-0.237) |
| 40-44 | 0.197 (0.195-0.198) | 0.206 (0.176-0.239) | 0.207 (0.173-0.246) | 0.208 (0.169-0.254) | 0.210 (0.165-0.264) | 0.211 (0.160-0.274) | 0.213 (0.155-0.286) | 0.215 (0.149-0.299) | 0.216 (0.144-0.313) | 0.218 (0.138-0.329) | 0.220 (0.132-0.345) | 0.222 (0.126-0.363) |
| 45-49 | 0.315 (0.314-0.317) | 0.338 (0.289-0.394) | 0.340 (0.283-0.405) | 0.342 (0.277-0.418) | 0.345 (0.270-0.433) | 0.347 (0.263-0.451) | 0.350 (0.254-0.471) | 0.353 (0.246-0.492) | 0.356 (0.237-0.516) | 0.360 (0.228-0.543) | 0.364 (0.219-0.571) | 0.368 (0.209-0.602) |
| 50-54 | 0.425 (0.423-0.426) | 0.455 (0.388-0.529) | 0.457 (0.381-0.544) | 0.460 (0.373-0.562) | 0.463 (0.363-0.582) | 0.466 (0.353-0.606) | 0.470 (0.341-0.632) | 0.474 (0.330-0.661) | 0.478 (0.318-0.693) | 0.483 (0.306-0.728) | 0.488 (0.293-0.767) | 0.494 (0.281-0.809) |
| 55-59 | 0.502 (0.500-0.504) | 0.532 (0.454-0.619) | 0.535 (0.446-0.636) | 0.537 (0.435-0.656) | 0.540 (0.424-0.679) | 0.544 (0.411-0.706) | 0.548 (0.398-0.736) | 0.552 (0.384-0.769) | 0.557 (0.370-0.806) | 0.562 (0.356-0.847) | 0.568 (0.341-0.891) | 0.574 (0.327-0.940) |
| 60-64 | 1.108 (1.105-1.111) | 1.312 (1.120-1.528) | 1.319 (1.100-1.570) | 1.326 (1.075-1.619) | 1.334 (1.046-1.677) | 1.341 (1.014-1.742) | 1.350 (0.981-1.814) | 1.359 (0.946-1.895) | 1.370 (0.910-1.984) | 1.381 (0.874-2.082) | 1.395 (0.838-2.190) | 1.409 (0.802-2.308) |
| 65-69 | 3.109 (3.103- | 3.634 (3.102 | 3.661 (3.052 | 3.690 (2.990 | 3.719 (2.916 | 3.747 (2.833 | 3.775 (2.743 | 3.804 (2.648 | 3.834 (2.549 | 3.867 (2.447 | 3.902 (2.345 | 3.940 (2.243 |

| | | | | | | | | | | | | |
|---|---|---|---|---|---|---|---|---|---|---|---|---|
| | 3.115) | -4.230) | -4.357) | -4.506) | -4.676) | -4.865) | -5.074) | -5.303) | -5.553) | -5.827) | -6.126) | -6.452) |
| 70-74 | 5.307 (5.298-5.316) | 5.732 (4.893-6.672) | 5.773 (4.812-6.869) | 5.818 (4.714-7.105) | 5.867 (4.601-7.377) | 5.920 (4.476-7.686) | 5.975 (4.342-8.030) | 6.035 (4.200-8.412) | 6.098 (4.053-8.832) | 6.163 (3.901-9.288) | 6.230 (3.745-9.782) | 6.298 (3.585-10.315) |
| 75-79 | 7.365 (7.351-7.378) | 7.305 (6.236-8.504) | 7.342 (6.120-8.736) | 7.387 (5.985-9.020) | 7.437 (5.832-9.351) | 7.493 (5.666-9.729) | 7.557 (5.491-10.156) | 7.628 (5.309-10.633) | 7.708 (5.124-11.164) | 7.796 (4.934-11.749) | 7.890 (4.743-12.389) | 7.992 (4.549-13.089) |
| 80-84 | 9.470 (9.449-9.490) | 8.564 (7.310-9.970) | 8.558 (7.133-10.183) | 8.570 (6.943-10.465) | 8.598 (6.742-10.811) | 8.639 (6.532-11.217) | 8.693 (6.316-11.683) | 8.757 (6.095-12.207) | 8.833 (5.871-12.793) | 8.919 (5.645-13.441) | 9.015 (5.419-14.155) | 9.122 (5.193-14.940) |
| 85-89 | 11.458 (11.423-11.494) | 9.637 (8.225-11.219) | 9.531 (7.944-11.341) | 9.447 (7.653-11.536) | 9.387 (7.360-11.804) | 9.352 (7.071-12.143) | 9.342 (6.788-12.556) | 9.357 (6.512-13.043) | 9.395 (6.244-13.607) | 9.452 (5.983-14.246) | 9.528 (5.727-14.961) | 9.620 (5.476-15.755) |
| 90-94 | 12.653 (12.576-12.730) | 10.578 (9.028-12.316) | 10.400 (8.668-12.376) | 10.234 (8.290-12.498) | 10.085 (7.907-12.682) | 9.955 (7.527-12.927) | 9.846 (7.153-13.234) | 9.761 (6.793-13.607) | 9.700 (6.447-14.050) | 9.666 (6.118-14.569) | 9.661 (5.806-15.170) | 9.684 (5.512-15.860) |
| ≥95 | 13.526 (13.321-13.732) | 11.551 (9.856-13.453) | 11.386 (9.487-13.553) | 11.216 (9.084-13.700) | 11.044 (8.657-13.890) | 10.872 (8.218-14.120) | 10.705 (7.776-14.391) | 10.549 (7.340-14.708) | 10.408 (6.917-15.077) | 10.287 (6.509-15.505) | 10.186 (6.121-15.997) | 10.109 (5.754-16.559) |
| ASR | 0.762 (0.762-0.763) | 0.768 (0.682-0.855) | 0.769 (0.660-0.878) | 0.771 (0.637-0.905) | 0.773 (0.612-0.934) | 0.775 (0.585-0.965) | 0.777 (0.556-0.998) | 0.780 (0.526-1.033) | 0.782 (0.494-1.071) | 0.785 (0.460-1.110) | 0.788 (0.425-1.151) | 0.791 (0.389-1.193) |

ASR=Annual Standardized Prevalence Ratio.

### H: Table S4 Prevalence of Heart Failure in Females in China (year 2021–2035, %)

| Age | 2021 | 2025 | 2026 | 2027 | 2028 | 2029 | 2030 | 2031 | 2032 | 2033 | 2034 | 2035 |
|---|---|---|---|---|---|---|---|---|---|---|---|---|
| 15-19 | 0.096 (0.095-0.097) | 0.099 (0.083-0.116) | 0.099 (0.081-0.119) | 0.099 (0.079-0.122) | 0.099 (0.076-0.126) | 0.100 (0.074-0.131) | 0.100 (0.071-0.136) | 0.100 (0.068-0.142) | 0.101 (0.065-0.148) | 0.101 (0.062-0.155) | 0.102 (0.059-0.163) | 0.103 (0.056-0.171) |
| 20-24 | 0.093 (0.092-0.094) | 0.098 (0.083-0.116) | 0.099 (0.081-0.119) | 0.099 (0.079-0.122) | 0.100 (0.077-0.127) | 0.100 (0.074-0.131) | 0.100 (0.071-0.137) | 0.101 (0.068-0.143) | 0.101 (0.066-0.149) | 0.102 (0.063-0.156) | 0.102 (0.060-0.164) | 0.103 (0.057-0.172) |
| 25-29 | 0.091 (0.090-0.092) | 0.098 (0.083-0.115) | 0.099 (0.081-0.118) | 0.099 (0.079-0.122) | 0.100 (0.077-0.127) | 0.100 (0.074-0.132) | 0.101 (0.072-0.137) | 0.102 (0.069-0.144) | 0.102 (0.066-0.150) | 0.103 (0.063-0.158) | 0.104 (0.060-0.166) | 0.105 (0.057-0.174) |
| 30-34 | 0.095 (0.094-0.096) | 0.104 (0.087-0.122) | 0.104 (0.086-0.126) | 0.105 (0.084-0.130) | 0.106 (0.082-0.135) | 0.107 (0.079-0.140) | 0.108 (0.077-0.147) | 0.109 (0.074-0.153) | 0.110 (0.071-0.161) | 0.111 (0.068-0.169) | 0.112 (0.065-0.178) | 0.113 (0.062-0.188) |
| 35-39 | 0.121 (0.120-0.122) | 0.133 (0.112-0.157) | 0.135 (0.111-0.162) | 0.136 (0.108-0.168) | 0.137 (0.106-0.174) | 0.138 (0.103-0.182) | 0.140 (0.099-0.190) | 0.141 (0.096-0.199) | 0.143 (0.092-0.209) | 0.144 (0.089-0.221) | 0.146 (0.085-0.233) | 0.148 (0.081-0.246) |
| 40-44 | 0.174 (0.173-0.175) | 0.192 (0.162-0.226) | 0.194 (0.160-0.233) | 0.196 (0.156-0.242) | 0.198 (0.153-0.252) | 0.200 (0.149-0.263) | 0.203 (0.144-0.276) | 0.205 (0.139-0.290) | 0.208 (0.134-0.305) | 0.210 (0.129-0.322) | 0.213 (0.124-0.340) | 0.216 (0.118-0.359) |
| 45-49 | 0.260 (0.258-0.261) | 0.288 (0.242-0.338) | 0.290 (0.239-0.349) | 0.293 (0.234-0.362) | 0.296 (0.228-0.377) | 0.300 (0.222-0.394) | 0.304 (0.216-0.413) | 0.307 (0.209-0.434) | 0.312 (0.201-0.457) | 0.316 (0.194-0.483) | 0.321 (0.186-0.511) | 0.326 (0.179-0.542) |
| 50-54 | 0.340 (0.339-0.342) | 0.374 (0.315-0.439) | 0.377 (0.310-0.453) | 0.380 (0.303-0.469) | 0.384 (0.296-0.488) | 0.388 (0.288-0.510) | 0.393 (0.279-0.534) | 0.397 (0.270-0.561) | 0.403 (0.260-0.591) | 0.408 (0.251-0.624) | 0.414 (0.241-0.661) | 0.421 (0.231-0.701) |
| 55-59 | 0.399 (0.397-0.400) | 0.434 (0.365-0.510) | 0.437 (0.359-0.526) | 0.441 (0.352-0.544) | 0.446 (0.343-0.566) | 0.450 (0.334-0.591) | 0.455 (0.323-0.618) | 0.460 (0.312-0.649) | 0.466 (0.301-0.683) | 0.472 (0.289-0.721) | 0.478 (0.278-0.763) | 0.485 (0.266-0.808) |
| 60-64 | 0.840 (0.837-0.843) | 1.006 (0.848-1.183) | 1.017 (0.836-1.223) | 1.028 (0.820-1.268) | 1.039 (0.800-1.320) | 1.050 (0.778-1.379) | 1.062 (0.754-1.444) | 1.074 (0.729-1.515) | 1.086 (0.702-1.595) | 1.100 (0.675-1.682) | 1.115 (0.648-1.778) | 1.131 (0.620-1.883) |
| 65-69 | 2.360 (2.355-2.364) | 2.772 (2.336-3.259) | 2.811 (2.311-3.379) | 2.851 (2.274-3.517) | 2.891 (2.227-3.674) | 2.931 (2.172-3.848) | 2.970 (2.110-4.039) | 3.009 (2.043-4.248) | 3.050 (1.972-4.477) | 3.092 (1.898-4.727) | 3.136 (1.822-5.000) | 3.182 (1.745-5.297) |
| 70-74 | 4.087 (4.080-4.095) | 4.383 (3.695-5.153) | 4.447 (3.655-5.345) | 4.515 (3.601-5.570) | 4.586 (3.533-5.828) | 4.660 (3.454-6.118) | 4.737 (3.366-6.441) | 4.816 (3.269-6.798) | 4.898 (3.167-7.190) | 4.983 (3.058-7.618) | 5.068 (2.945-8.081) | 5.155 (2.827-8.582) |
| 75-79 | 5.725 (5.713-5.736) | 5.550 (4.678-6.525) | 5.615 (4.616-6.749) | 5.689 (4.537-7.018) | 5.769 (4.444-7.331) | 5.856 (4.340-7.688) | 5.950 (4.227-8.091) | 6.051 (4.108-8.542) | 6.161 (3.983-9.044) | 6.278 (3.853-9.599) | 6.401 (3.719-10.207) | 6.530 (3.582-10.872) |
| 80 | 7.831 | 6.955 | 6.989 | 7.039 | 7.105 | 7.184 | 7.276 | 7.381 | 7.498 | 7.628 | 7.769 | 7.923 |

| | | | | | | | | | | | | |
|---|---|---|---|---|---|---|---|---|---|---|---|---|
| -84 | (7.814-7.847) | (5.862-8.177) | (5.745-8.400) | (5.614-8.684) | (5.473-9.029) | (5.325-9.432) | (5.170-9.895) | (5.010-10.419) | (4.847-11.007) | (4.682-11.662) | (4.514-12.388) | (4.345-13.191) |
| 85-89 | 10.831 (10.804-10.857) | 9.080 (7.653-10.676) | 9.024 (7.417-10.846) | 8.989 (7.169-11.090) | 8.979 (6.917-11.411) | 8.995 (6.666-11.810) | 9.036 (6.420-12.289) | 9.103 (6.179-12.850) | 9.194 (5.943-13.497) | 9.309 (5.713-14.233) | 9.445 (5.488-15.061) | 9.602 (5.266-15.986) |
| 90-94 | 13.127 (13.078-13.175) | 10.845 (9.140-12.752) | 10.705 (8.798-12.867) | 10.579 (8.436-13.052) | 10.471 (8.065-13.307) | 10.383 (7.695-13.634) | 10.319 (7.331-14.034) | 10.281 (6.978-14.514) | 10.271 (6.639-15.078) | 10.291 (6.316-15.736) | 10.344 (6.010-16.496) | 10.431 (5.720-17.367) |
| ≥95 | 14.703 (14.598-14.807) | 12.128 (10.219-14.264) | 11.992 (9.854-14.417) | 11.852 (9.449-14.626) | 11.712 (9.020-14.888) | 11.575 (8.577-15.201) | 11.444 (8.129-15.566) | 11.325 (7.686-15.990) | 11.223 (7.254-16.478) | 11.143 (6.838-17.039) | 11.088 (6.441-17.683) | 11.061 (6.065-18.418) |
| ASR | 0.628 (0.628-0.629) | 0.629 (0.554-0.703) | 0.633 (0.538-0.727) | 0.637 (0.520-0.753) | 0.642 (0.501-0.782) | 0.646 (0.480-0.813) | 0.651 (0.457-0.846) | 0.657 (0.432-0.881) | 0.662 (0.406-0.919) | 0.668 (0.378-0.958) | 0.674 (0.349-0.999) | 0.679 (0.317-1.042) |

ASR=Annual Standardized Prevalence Ratio.

**I: Table S5 Age-specific number of Heart Failure cases across different age groups in different years**

| Age group, years | 2021 | 2025 | 2035 |
|---|---|---|---|
| 0-4 | 229653.9 (191419.98-271092.01) | 158353.83 (122859.21-193848.45) | 152934.01 (35467-270401.02) |
| 5-9 | 172002.8 (120944.6-233516.96) | 145242.92 (96823.26-193662.57) | 94700.32 (0-215322.03) |
| 10-14 | 97360.69 (75682.11-121377.42) | 99130.31 (41078.82-157181.79) | 61114.98 (0-219429.71) |
| 15-19 | 76281.82 (57469.94-97786.05) | 87248.33 (31711.55-142785.11) | 84796.6 (0-342464.9) |
| 20-24 | 72261.29 (57196.68-88953.46) | 80567.31 (28967.73-132166.88) | 93851.57 (0-382982.1) |
| 25-29 | 83032.21 (62984.62-106152.23) | 82815.59 (28806.09-136825.08) | 88660.46 (0-360232.07) |
| 30-34 | 120425.26 (83570.5-164556.34) | 104487.08 (39774.35-169199.82) | 87864.13 (0-336851.58) |
| 35-39 | 134333.92 (99184.53-171592.31) | 165337.81 (86554.07-244121.55) | 119027.94 (0-369067.3) |
| 40-44 | 169778.94 (118860.27-228362.81) | 207695.21 (139814.81-275575.6) | 214468.9 (0-505636.1) |
| 45-49 | 317763.99 (247752.9-397698.76) | 290707.09 (223572.53-357841.65) | 419410.32 (45645.58-793175.06) |
| 50-54 | 462965.96 (360732.22-615299.16) | 478660.55 (383199.16-574121.94) | 468048.49 (121891.58-814205.4) |
| 55-59 | 494914.45 (361210.75-667069.81) | 565239.07 (458872.78-671605.37) | 475980.14 (149508.61-802451.67) |
| 60-64 | 711347.05 (573774.13-864213.39) | 1093513.75 (914681.35-1272346.16) | 1391177.45 (607818.96-2174535.94) |
| 65-69 | 2092602.56 (1686361.17-2558920.43) | 2248427.97 (1887842.27-2609013.67) | 3821986.86 (1725606.68-5918367.04) |
| 70-74 | 2493753.44 (1953403.24-3112149.16) | 3283364.17 (2756750.61-3809977.72) | 4702124.57 (2124100.58-7280148.56) |
| 75-79 | 2151831 (1670994.62-2717410.62) | 2501846.93 (2100340.97-2903352.88) | 4009099.17 (1809196.37-6209001.97) |
| 80-84 | 1692066.6 (1384371.68-2096131.69) | 1651635.54 (1385781.96-1917489.12) | 3714846.8 (1672133.19-5757560.42) |
| 85-89 | 1053620.26 (858679.72-1295688.66) | 1052842.24 (882365.53-1223318.95) | 1884757.21 (845238.67-2924275.75) |
| 90-94 | 381112.51 (289793.05-494911.9) | 456177.27 (381805.48-530549.06) | 643663.79 (287467.01-999860.56) |
| ≥95 | 92617.94 (66074.44-129518.54) | 104102.3 (86959.87-121244.73) | 167823.02 (74502.17-261143.88) |

**J: Table S6 Heart failure treatment cost per capita (2017 INT$)**

| Age group, years | Male | Female |
|---|---|---|
| 15-19 | 2515.829 (1076.771-4857.952) | 2311.820 (779.701-3125.863) |
| 20-24 | 2515.829 (1076.771-4857.952) | 2311.820 (779.701-3125.863) |
| 25-29 | 2515.829 (1076.771-4857.952) | 2311.820 (779.701-3125.863) |
| 30-34 | 2637.633 (1948.290-2956.346) | 2238.629 (953.429-2806.493) |
| 35-39 | 2571.786 (2114.786-3541.798) | 2421.140 (1333.369-2788.182) |
| 40-44 | 2682.079 (2236.495-3492.191) | 2628.427 (1615.473-3124.873) |
| 45-49 | 2851.943 (2355.750-3943.028) | 2512.620 (2159.953-3984.858) |
| 50-54 | 3286.758 (2608.978-5188.255) | 2517.421 (2161.002-4865.515) |
| 55-59 | 3851.943 (3068.937-7497.100) | 2897.489 (2509.708-5914.320) |
| 60-64 | 4402.177 (3358.696-8928.951) | 3499.409 (3210.987-7432.978) |
| 65-69 | 5343.961 (4215.684-10224.311) | 4352.872 (3774.387-9947.769) |
| 70-74 | 6099.156 (5259.225-9882.422) | 5207.016 (4534.780-9918.090) |
| 75-79 | 7084.377 (5342.309-11463.360) | 6179.463 (5317.930-11531.145) |
| 80-84 | 7803.200 (6063.041-12350.262) | 7341.848 (5578.108-12735.883) |
| 85-89 | 8529.589 (6016.547-11350.710) | 8402.634 (5320.305-11313.725) |
| 90-94 | 8654.335 (5761.395-11069.264) | 9316.168 (4963.876-11856.257) |
| 95-99 | 8278.621 (5527.340-9314.985) | 8092.634 (3789.100-12416.460) |
| ≥100 | 10614.733 (2721.092-15855.898) | 9182.279 (4859.647-12184.794) |

**K: Table S7 Sensitivity Analysis of the Macroeconomic Burden of Heart Failure in China Under Varying Discount Rates (2025–2035)**

| | **discounted at 0% (lower and upper bound*)** | **discounted at 1% (lower and upper bound*)** | **discounted at 2% (lower and upper bound*)** | **discounted at 4% (lower and upper bound*)** | **discounted at 5% (lower and upper bound*)** |
|---|---|---|---|---|---|
| Economic Loss (Billions of 2017 INT$) | 1209.250 (873.787, 1644.584) | 1136.823 (823.534, 1542.669) | 1070.307 (777.315,1449.193) | 952.822 (695.497,1284.401) | 900.899 (659.254,1211.715) |
| Proportion of GDP (%) | 0.265 (0.191, 0.360) | 0.263 (0.190,0.356) | 0.260 (0.189,0.353) | 0.256 (0.187,0.345) | 0.254 (0.186,0.341) |

*Uncertainty intervals in parentheses are calculated based on the lower and upper bounds of 95% uncertainty intervals for Projected HF Prevalence by Age Group in China derived from the Global Burden of Disease (GBD) Study 2021, adjusted for China-specific demographic and risk factor trends

**L: Table S8 Sensitivity Analysis of Projected Macroeconomic Burden of Heart Failure in China (2025–2035)**

| Year | Low Cost | | High Cost | |
|---|---|---|---|---|
| | Economic Loss, Billions of 2017 INT$ | Proportion of GDP, % | Economic Loss, Billions of 2017 INT$ | Proportion of GDP, % |
| 2025 | 46.552 | 0.137 | 46.552 | 0.137 |
| 2026 | 56.823 | 0.16 | 60.147 | 0.17 |
| 2027 | 67.241 | 0.183 | 73.865 | 0.201 |
| 2028 | 77.788 | 0.205 | 87.707 | 0.231 |
| 2029 | 88.567 | 0.226 | 101.825 | 0.259 |
| 2030 | 100.766 | 0.246 | 117.578 | 0.287 |
| 2031 | 113.798 | 0.267 | 134.284 | 0.315 |
| 2032 | 127.707 | 0.287 | 151.988 | 0.342 |
| 2033 | 142.486 | 0.308 | 170.692 | 0.368 |
| 2034 | 158.173 | 0.328 | 190.456 | 0.394 |
| 2035 | 175.033 | 0.348 | 211.543 | 0.42 |
| Total (Discounted) | 964.988 | 0.247 | 1120.893 | 0.287 |

GDP = gross domestic product.
Economic loss is expressed in billions of 2017 international dollars (2017INT$).
The total economic burden includes direct healthcare costs (e.g., hospitalizations, medications) and indirect costs (e.g., lost productivity, disability).

**References**

1 Chen S, Kuhn M, Prettner K, et al. The global economic burden of chronic obstructive pulmonary disease for 204 countries and territories in 2020-50: a health-augmented macroeconomic modelling study. *Lancet Glob Health* 2023; **11**(8): e1183-93.

2 Bloom DE, Chen S, Kuhn M, Mcgovern ME, Oxley L, Prettner K. The economic burden of chronic diseases: estimates and projections for china, japan, and south korea. *The Journal of the Economics of Ageing* 2020; **17**: 100163.

3 Chen S, Kuhn M, Prettner K, Bloom DE. The global macroeconomic burden of road injuries: estimates and projections for 166 countries. *Lancet Planet Health* 2019; **3**(9): e390-8.

4 Chen S, Cao Z, Prettner K, et al. Estimates and projections of the global economic cost of 29 cancers in 204 countries and territories from 2020 to 2050. *JAMA Oncol* 2023; **9**(4): 465-72.

5 Chen S, Kuhn M, Prettner K, Bloom DE. Noncommunicable diseases attributable to tobacco use in china: macroeconomic burden and tobacco control policies. *Health Aff (Millwood)* 2019; **38**(11): 1832-9.

6 Chen S, Bloom DE. The macroeconomic burden of noncommunicable diseases associated with air pollution in china. *PLoS One* 2019; **14**(4): e0215663.

7 Lucas RE. On the mechanics of economic development. *J Monet Econ* 1988; **22**(1): 3-42.

8 Abegunde D, Stanciole A. An estimation of the economic impact of chronic noncommunicable disease in selected countries. *World Health Organization, Department of Chronic Diseases and Health Promotion 2006* 2006.

9 Solow RM. A contribution to the theory of economic growth. *Q J Econ* 1956; **70**: 65-94.

10 World economic outlook database, october 2024. https://www.imf.org/en/Publications/WEO/weo-database/2024/October (accessed 2025-3-4, 2025).

11 Mincer JA. Schooling, experience, and earnings. Human behavior & social institutions No. 2. New York: National Bureau of Economic Research Inc., 1974.

12 Chen S, Kuhn M, Prettner K, Bloom DE. The macroeconomic burden of noncommunicable diseases in the united states: estimates and projections. *PLoS One* 2018; **13**(11): e0206702.

13 Wang H, Li Y, Chai K, et al. Mortality in patients admitted to hospital with heart failure in china: a nationwide cardiovascular association database-heart failure centre registry cohort study. *Lancet Glob Health* 2024; **12**(4): e611-22.

14 Barro RJ, Lee JW. A new data set of educational attainment in the world, 1950–2010. *J Dev Econ* 2013; **104**: 184-98.

15 GBD 2021 forecasting collaborators. Burden of disease scenarios for 204 countries and territories, 2022-2050: a forecasting analysis for the global burden of disease study 2021. *Lancet* 2024; **403**(10440): 2204-56.

16 Pwt 10.01. https://www.rug.nl/ggdc/productivity/pwt/ (accessed 2025-3-4, 2025).

17 ILO modelled estimates (ILOEST database) - ILOSTAT. https://ilostat.ilo.org/methods/concepts-and-definitions/ilo-modelled-estimates/ (accessed 2025-3-4, 2025).

18 World population prospects. https://population.un.org/wpp/ (accessed 2025-3-4, 2025).

19 World development indicators | DataBank. https://databank.worldbank.org/reports.aspx?source=2&series=NY.GNS.ICTR.ZS&country=# (accessed 2025-3-4, 2025).

20 Wang H, Chai K, Du M, et al. Prevalence and incidence of heart failure among urban patients in china: a national population-based analysis. Circ Heart Fail 2021; 14(10): e008406.

21 Neumann PJ, Ganiats TG, Russell LB, Sanders GD, Siegel JE. Cost-effectiveness in health and medicine. Oxford University Press, 2016.

22 Grossmann V, Steger T, Trimborn T. Dynamically optimal r&d subsidization*.* *Journal of Economic Dynamics and Control* 2013; **37**(3): 516-34.

23 Psacharopoulos G, Patrinos HA. Returns to investment in education: a decennial review of the global literature*.* *Educ Econ* 2018; **26**(5): 445-58.

24 Heckman JJ, Lochner LJ, Todd PE. Chapter 7 earnings functions, rates of return and treatment effects: the mincer equation and beyond. In: Hanushek E, Welch F, eds. Handbook of the Economics of EducationElsevier, 2006: 307-458.
